# Estimating behavioural relaxation induced by COVID-19 vaccines in the first months of their rollout

**DOI:** 10.1101/2024.11.07.24316893

**Authors:** Yuhan Li, Nicolò Gozzi, Nicola Perra

## Abstract

The initial rollout of COVID-19 vaccines has been challenged by logistical issues, limited availability of doses, scarce healthcare capacity, spotty acceptance, and variants of concern. Non-pharmaceutical interventions (NPIs) have been critical to support these phases. However, vaccines may have prompted behavioural relaxation, potentially reducing NPIs adherence. Epidemic models have explored this phenomenon, but they have not been validated against data. Moreover, recent surveys provide conflicting results on the matter. The extent of behavioural relaxation induced by COVID-19 vaccines is still unclear. Here, we aim to study this phenomenon in four regions. We implement five realistic epidemic models which include age structure, multiple virus strains, NPIs, and vaccinations. One of the models acts as a baseline, while the others extend it including different behavioural relaxation mechanisms. First, we calibrate the baseline model and run counterfactual scenarios to quantify the impact of vaccinations and NPIs. Our results confirm the critical role of both in reducing infection and mortality rates. Second, we calibrate the behavioural models and compare them to each other and to the baseline using different metrics. Including behavioural relaxation leads to a better fit of weekly deaths in three regions. However, the improvements are limited to a 2 − 10% reduction in weighted mean absolute percentage errors and these gains are generally offset by models’ increased complexity. Overall, we do not find clear signs of behavioural relaxation induced by COVID-19 vaccines on weekly deaths. Furthermore, our results suggest that if this phenomenon occurred, it generally involved only a firm minority of the population. Our work contributes to the retrospective validation of epidemic models developed amid the COVID-19 Pandemic and underscores the issue of non-identifiability of complex social mechanisms.

## Introduction

In the Global North the first dose of COVID-19 vaccines was administered on December 8, 2020 [1]. Vaccines led to a significant reduction in mortality and transmission rates [2–6]. Especially in the first months of their rollout, however, vaccination efforts have encountered many challenges due to logistical issues, limited stockpiles and healthcare capacity, vaccine nationalism, and spotty acceptance [7]. A study from the United States, for instance, showed that counties with limited healthcare resources were also more likely to achieve lower COVID-19 vaccination rates [8]. Vaccine nationalism and socioeconomic inequities led to a concentration of doses in high-income countries [9, 10]. The insufficient vaccination coverage in many areas proved inadequate to prevent subsequent waves, thus leading to increased disease burden and to the implementation of additional interventions to curb the spread of SARS-CoV-2 [6, 11]. On a global scale, initial vaccine acceptance varied significantly across different regions, ranging from 13% in Iraq to 97% in Vietnam according to surveys conducted before the start of vaccine rollout [12]. Furthermore, the effectiveness of vaccines - especially in preventing transmission - was challenged by the emergence of new variants of concern (VOC) such as Alpha and Delta [13, 14].

While non-pharmaceutical interventions (NPIs) have been key to support vaccination efforts during these initial phases [5, 6, 15], their adoption is shaped by many factors such as perceived susceptibility, severity, barriers to actions, exposure to (mis)information, and peer-effects [16–22]. It is natural to wonder whether the arrival of COVID-19 vaccines impacted the adoption of NPIs. Indeed, the start of vaccinations might have lowered the perception of risk at least in some groups of the population, which in turn might have led to a relaxation of NPIs. The potential effects of this phenomenon, which for simplicity we will refer to as <u>behavioural relaxation</u>, have been explored via epidemic models in realistic, yet theoretical, scenarios during the first months of the vaccine rollout [23–26]. The results from these efforts suggest how behavioural relaxation could reduce the positive gains brought about by vaccines thus leading to higher disease burden. The empirical evidence does not provide a clear picture of the extent of behavioural relaxation. Indeed, some survey data report that vaccinated individuals had 1.31 times more social contacts [27] and increased mobility levels [28] with respect to non-vaccinated people. A regression analysis conducted considering different mobility data in London shows a positive association between mobility and vaccinations [28]. Other surveys conducted in Brazil, Italy, South Africa, and the United Kingdom, indicate that public transport usage increased by up to 10% after the rollout of first dose of vaccine [29]. However, the results from large, and repeated, cross-sectional surveys conducted in France provide limited support for a systematic behavioural relaxation, especially during the initial phases of the vaccine rollout [30].

In this context, we investigate the interplay between vaccinations and NPIs during the first months of the COVID-19 vaccine rollout in four regions: British Columbia (Canada), Lombardy (Italy), London (United Kingdom), and São Paulo (Brazil). These regions have been selected to sample different epidemiological, socioeconomic, and socio-demographic contexts, as well as different vaccine rollout schedules and coverages. We set to quantify the extent of behavioural relaxations induced by the start of vaccination campaigns and estimate their potential impact on reported weekly deaths via epidemic models. Indeed, as mentioned above, the models published so far to capture such behavioural relaxation have not been validated against real data [23–26]. Besides, they have not been compared among them nor against a simpler baseline. By using the data collected and made available over the last years, we can address these gaps. To this end, we develop a series of stochastic compartmental epidemic models, integrating vaccinations, variants of concern, age-structure, NPIs, as well as individuals’ behavioural relaxation linked to vaccines. In particular, we consider a baseline model without behavioural relaxation mechanisms and four models that instead include them. In these, we introduce explicit compartments that account for non-compliant individuals who relaxed their protective behaviours as a result of the start of vaccinations. These models, which we will refer to as <u>behavioural models</u>, differ according to the mechanisms used to describe the transitions in and out of non-compliant behavioural compartments.

To set the stage, we first calibrate the baseline model to reported data (i.e., weekly deaths) in the four locations and run two counterfactual scenarios to quantify the impact of vaccines and of NPIs on COVID-19 burden. Our results clearly confirm the crucial role played by both in reducing deaths and infections. Then, we calibrate and compare the other four behavioural models against each other and the baseline. We find that all models provide similar outcomes and are able to reproduce the trajectory of the COVID-19 Pandemic in the four locations during the period considered. By studying the weighted mean absolute percentage errors (wMAPEs) we find that incorporating behavioural relaxation mechanisms improves model accuracy in three regions under study by 2 − 10%. However, when accounting for the increased complexity of these models via the Akaike and Bayesian Information Criterion (AIC and BIC) [31, 32], we find the baseline to be the most likely model in three out of the four locations according to the AIC. Indeed, only in Lombardy one of the behavioural models is selected as the most likely. According to the BIC instead, the baseline is the most likely model in all locations. This metric is known to favour simpler models with respect to the AIC. Nevertheless, we find how in Lombardy the baseline model is only marginally more likely than one of the behavioural models.

Overall, by applying the Occam’s razor principle, our results do not support the inclusion of behavioural relaxation mechanisms across all regions. It is important to notice, however, how our findings do not exclude, as suggested by several surveys, that some people did in fact relaxed their behaviours as result of the start of the vaccination campaign. Indeed, the phase space selected in the calibration suggest that, if behavioural relaxation took place, it was limited to a firm minority of the population thus not leaving clear marks on weekly reported COVID-19 deaths. Finally, our findings highlight the issue of non-identifiability of complex behavioural dynamics in epidemic models.

Our results confirm the critical role of vaccines and NPIs in mitigating deaths and infections during the initial phases of the COVID-19 mass vaccination campaigns. At the same time, they estimate the extent of behavioural relaxation induced by vaccines in four regions. As we reflect on the COVID-19 Pandemic and prepare for the next one, studies that test and compare different models developed amid the emergency are of clear importance.

## Results

We implement and compare five epidemic models. The first acts as a baseline. The others build on it and include different behavioural relaxation mechanisms. The four behavioural models combine and extend approaches from the literature [23, 24]. To explore different epidemiological, socioeconomic and socio-demographic contexts, as well as different vaccine rollout schedules and coverages we consider four regions of the world: British Columbia (Canada), Lombardy (Italy), London (United Kingdom), and São Paulo (Brazil). All models are calibrated to confirmed weekly deaths via an Approximate Bayesian Computation-Sequential Monte Carlo (ABC-SMC) method [33]. While we provide a summary of the models in the next two sections, we refer the reader to the Material and Methods as well as section 1.1 in Supplementary Information for full details.

### Baseline model

The baseline model (<u>baseline</u>) is a Susceptible-Latent-Infected-Recovered (SLIR) epidemic model integrating vaccinations, NPIs, age-structured contact matrices, multiple virus strains, and disease-related deaths. It constitutes the core upon which the other four models are built. We include age-stratified vaccinations by using real data [34–38]. For simplicity, we assume a single dose regiment and ignore the time required to develop full protection after inoculation. Furthermore, we assume that only Susceptible individuals are eligible for vaccination. We estimate the impact of NPIs on social contacts using mobility data from the COVID-19 Community Mobility Report published by Google LLC [39] and the Oxford Coronavirus Government Response Tracker (OxCGRT) [40]. This data is used to modulate the contact matrices as function of time. We also consider the emergence and spread of a second virus strain. According to virological surveillance data, during the period under consideration, the Alpha variant emerged and replaced the ancestral type in British Columbia and Lombardy, while Delta replaced the Gamma VOC in São Paulo. London, during the time interval under investigation, faced primarily a wave dominated by the Alpha VOC [41–43]. In our models, we assume Alpha and Delta to have higher transmissibility [44, 45], vaccine-induced immunity escape potential [44, 46, 47], and shorter latent period with respect to previously circulating strains [48–52]. We refer the reader to the Materials and Methods for more details.

### Behavioural relaxation models

Building on the baseline and the literature, we explore four different behavioural relaxation models. To this end, we extend the compartmental structure of the baseline by introducing non-compliant (NC) compartments to account for susceptible individuals (both vaccinated or not) who relax their COVID-19 safe behaviours as result of the vaccine rollout. Individuals in the NC compartments have a probability of infection that is *r* (*r* > 1) times higher than that of compliant individuals [23, 24, 27]. The four behavioural models differ for the mechanisms regulating the transitions from and to compliance. Following Ref. [23] in the first (<u>model 1</u>) and second (<u>model 2</u>) behavioural model we assume that all susceptible individuals (vaccinated or not) might enter/leave the NC compartments. In detail, in <u>model 1</u> the transition rates between compliance and non-compliance are constant and equal to *α* and *γ* respectively. In <u>model 2</u>, susceptible individuals enter/leave the NC compartment at varying rates. The transition rate to non-compliant behaviour is set as a function of the fraction of vaccinated individuals and a parameter *α*. The reverse transition rate is instead set as a function of the number of reported daily deaths per 100, 000 and a parameter *γ*. Indeed, daily deaths have been often used, especially by media, to characterize the status of the Pandemic and are known to have influenced individuals’ adherence NPIs [16]. Following Ref. [24] the third (<u>model 3</u>) and fourth (<u>model 4</u>) behavioural models are respectively analogous to model 1 and model 2. However, in these two, only susceptible vaccinated individuals might transition to non-compliant compartments.

### Vaccines rollout, epidemic progression and NPIs in the four regions under study

The rollout of COVID-19 vaccines is a key part of our work. Hence, we start by providing some information about the initial phases of vaccinations in the four regions under study. In Fig. 1-A, we show the 7-day moving average of the fraction of daily newly vaccinated individuals across all age groups (shaded areas) and in the 70+ age group (solid lines) from the start of the vaccination rollout until the end date of the period under consideration. COVID-19 vaccination campaigns started on 2020*/*12*/*19 in British Columbia, 2020*/*12*/*08 in London, 2020*/*12*/*27 in Lombardy, and 2021*/*01*/*18 in São Paulo [34–38]. In all locations, we observe a peak in the first month during which the initial doses were mainly administered to healthcare workers and fragile individuals. A similar behaviour can be seen for the 70+ age group. Moreover, the vaccine rates of this group show a second peak earlier with respect to the overall vaccination rates in the four regions, reflecting the priority given to the elderly population. Additionally, we observe how the vaccination rate in London was concentrated during the second to the fifth month since the rollout started. In British Columbia and Lombardy instead, vaccination started on a wider scale (i.e., beyond the prioritization of fragile individuals and healthcare workers) from the third month of the rollout, and even later in São Paulo (from the fourth month).

**Fig 1.**
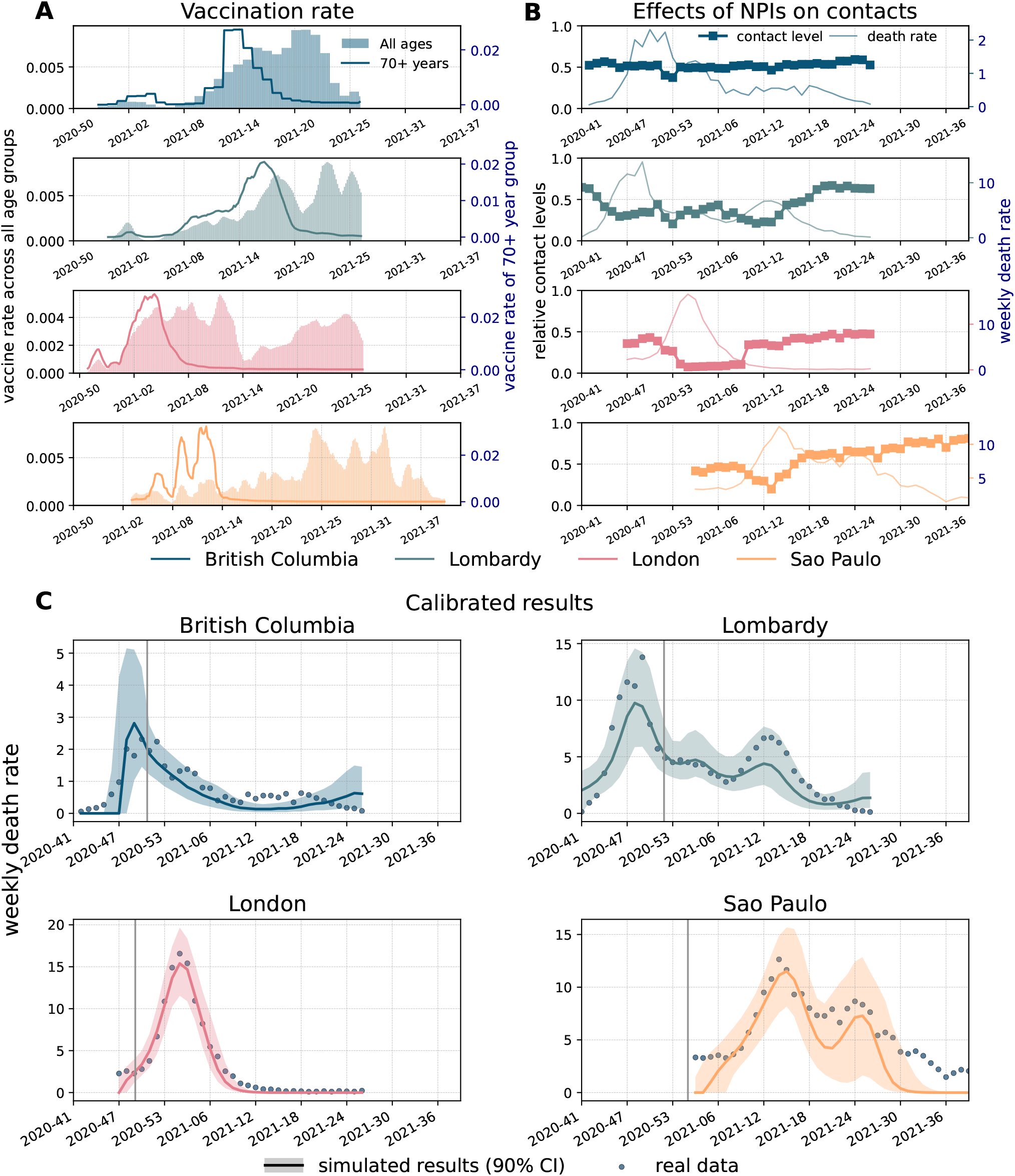
Weekly deaths, vaccinations, contacts reduction, and calibration of baseline model. The x-axis indicates the year and week. A) Fraction of daily newly vaccinated individuals across all age groups (shaded areas) and within the 70+ age-group (solid lines) in the four regions from up to down: British Columbia, Lombardy, London, and São Paulo. B) Contact levels during the Pandemic with respect to a pre-pandemic baseline. C) Reported data describing weekly deaths per 100, 000 (dots) and the results from the calibrated baseline model (solid lines representing the medians, shaded areas the 90% confidence intervals). The grey vertical lines mark the start of vaccinations in different regions.

To better understand the epidemic contexts in the periods under examination, next we discuss the evolution of the Pandemic and of NPIs in the four regions. In Fig. 1-B we show the confirmed deaths per 100, 000 (solid lines). We can observe differences in the timing, shape, and intensity of the Pandemic across the four regions. Indeed, within the time horizon of interest, British Columbia and London experienced a single peak concentrated around the end of 2020. The peak in London was particularly intense and fuelled by the spread of the Alpha VOC which was initially detected there and traced back to a set of transmission chains that occurred in September 2020 [53]. In British Columbia, instead, the peak was more than five times less intense and followed by a slower decrease with fluctuations due to the Alpha variant replacing the wild type [54]. In Lombardy, we observe an intense peak (comparable to London) right before the end of the year and a second, less pronounced, peak in early April mainly due to the lifting of some of the NPIs and the spreading of Alpha [5]. Also in São Paulo we observe two intense peaks, which however are much closer both in terms of intensity and timing. Genomic data in this region suggests that the two peaks were driven by the rapid spread of the Gamma VOC first followed by the arrival and spread of the Delta VOC [43, 55, 56]. We note how the first of these two peaks take place in April, hence months later than the main peak in the other three areas here under examination.

In Fig. 1-B we also show the effect of NPIs on contacts estimated from the COVID-19 Community Mobility Report released by Google [39] and the Oxford Coronavirus Government Response Tracker (OxCGRT) [40]. We use this data to compute the contact levels during the Pandemic with respect to a pre-Pandemic baseline. Indeed, this data has been often used as a proxy of NPIs adoption, especially during the first two years of the Pandemic [16]. We refer the reader to the Materials and Methods for details. The plot suggests that, among the regions considered, individuals in London adopted the strictest NPIs. This is likely due to the emergence and rapid spread of the Alpha VOC that resulted in strong social distancing policies. These measures led to a significant reduction in contacts ranging from 10% to 50% with respect to the pre-pandemic contact levels. A similar trend, though not as strong, is observed in Lombardy where contacts rapidly dropped as the 2020 winter season progressed. In British Columbia, despite a visible drop at the end of 2020, we observe a slow increasing trend centred around 50% with respect to the pre-pandemic baseline. Similar trends are observed in São Paulo where, however, we can observe a much steeper increase in contacts, back to, and even larger than, the pre-pandemic baseline in the second half of 2021.

### Baseline calibration

In Fig. 1-C, we show the weekly deaths as reported by official surveillance and as estimated by the baseline epidemic model in the four regions considered. In the figure, we plot medians along with 90% confidence intervals (CI) computed considering 1000 stochastic trajectories sampled from the joint posterior distribution estimated via the ABC-SMC calibration (see Materials and Methods for details). We highlight the start of the vaccination campaign in each location with vertical solid lines. To account for local differences in the epidemic trajectory, the starting point of our simulations is left as a free, calibrated, parameter (see Materials and Methods for more details). Furthermore, the simulation horizons in the four regions are set to capture the local epidemic wave(s) in the first months of the vaccine rollout. More precisely, we run simulations until 2021/07/04 in British Columbia, London, and Lombardy. In case of São Paulo we run until 2021/10/03 to capture the late waves of infection experienced there with respect to the other regions. Interestingly, most reported data points fall within the 90% CI of the calibrated baseline, which suggests the effectiveness of the model in fitting the unfolding of the Pandemic in the four locations despite the differences in terms of shapes and intensities of each epidemic curve. The analysis of the posterior distributions of free parameters, shown in section 5.3 of the Supplementary Information, indicates London as the region with the highest basic reproductive number *R*_0_. This is likely due the dominance of the Alpha VOC in London at the start of our simulation window.

### Estimating the impact of vaccines and NPIs via counterfactual scenarios

To estimate the impact of vaccines on COVID-19 deaths and infections we run a counterfactual scenario where they are removed from the baseline model. To this end, we first calibrate the model in the four locations. Then, we run matched simulations where we remove vaccinations. Hence, we quantify the effect of vaccines by computing the relative deaths difference (RDD) between the total number of deaths in a model with vaccines and those observed in an equivalent model without vaccines (see more details in Materials and Methods). The RDD for vaccines is shown in Fig. 2-A. The median values of RDD are greater than zero highlighting the clear positive effects of vaccinations. In particular, we find an RDD of 10.33% (90% CI: [−2.50%,25.01%]) in British Columbia, 15.90% ([9.65%,23.99%]) in Lombardy, 1.20% ([−0.06%,4.82%]) in London, and 50.69% ([45.77%,56.16%]) in São Paulo. The difference in RDD across the four regions is possibly due to several factors including timing and coverage of vaccines, local epidemiological context (e.g., VOC circulating), and NPIs in place. Notably, São Paulo, which exhibits the highest RDD, achieved also the highest vaccine coverage (78% of the population) by the end date of the simulation window. This is significantly larger than the coverage in British Columbia (69%), Lombardy (63%), and London (61%). As noted above, São Paulo is the region with the lowest reduction of contacts due to NPIs. This might contribute to enhance the role of vaccines. Notably, the median value of RDD in São Paulo corresponds to more than 70*K* additional deaths averted (see section 3.5 in the Supplementary Information). Though British Columbia has marginally higher vaccine coverage compared to Lombardy, its RDD is however the second lowest. This discrepancy can be explained by the relatively lower and slower epidemic progression in this region. As mentioned, this location experienced, at the peak, a burden of the disease about five times lower than the other three. The RDD values in this region are reflective of a very small absolute difference of deaths between the two scenarios (144 deaths as shown in section 3.5 in Supplementary Information). The RDD in London is the lowest value. In London, the early 2021 wave was fuelled by the Alpha VOC which was more transmissible and able to reduce the vaccines’ protection from infection. Besides, according to Fig. 1-A, wide-scale vaccination started from week 1 of 2021 when the number of deaths nearly reached the peak. Additionally, as shown above, London faced the strictest NPIs among the regions considered bringing contacts down even to 10% of pre-Pandemic levels. Finally, as reported above, the vaccine coverage in London by the end of our simulation window, is the smallest in the group of countries under investigation.

**Fig 2.**
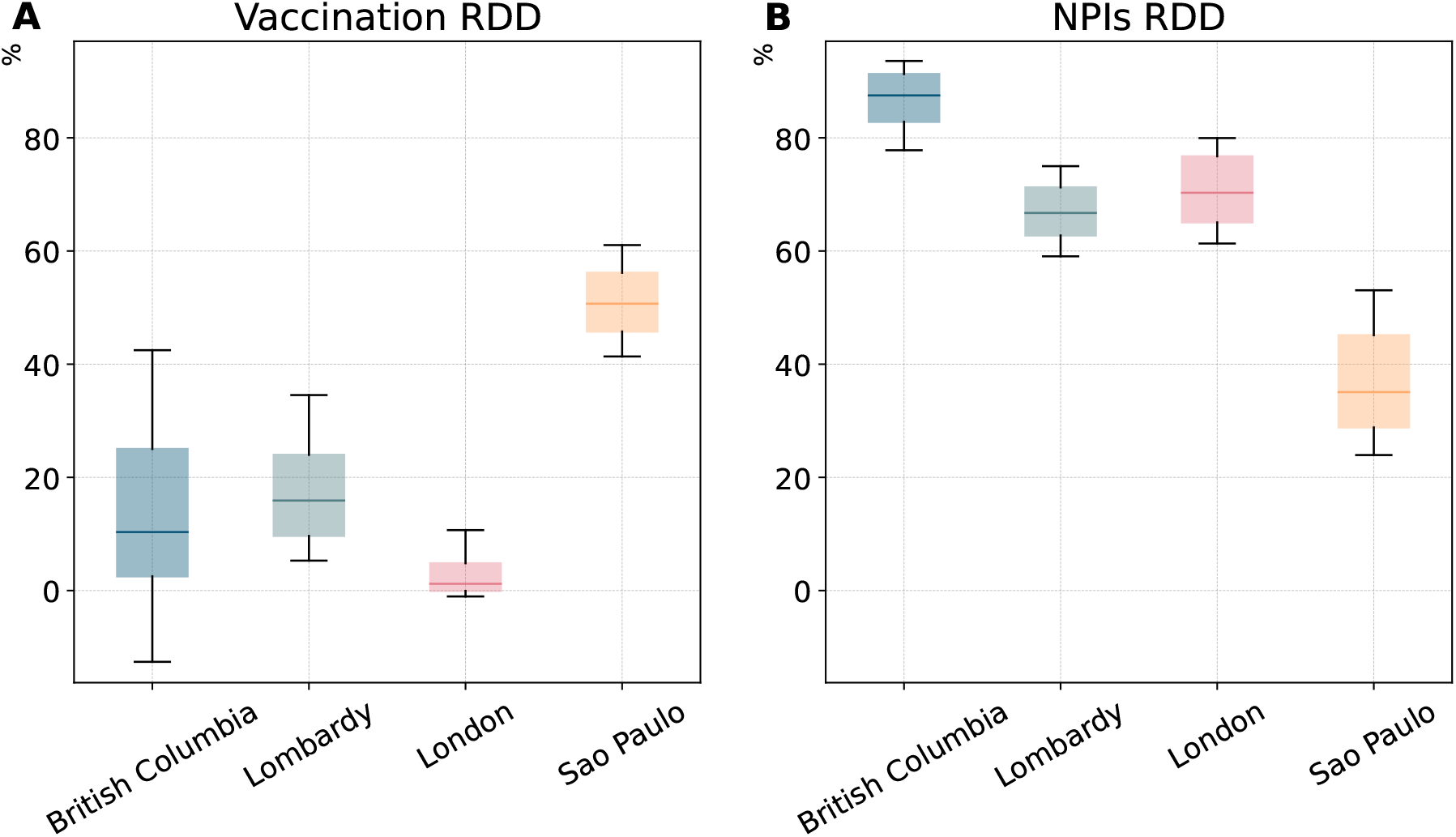
Impact of vaccines and non-pharmaceutical interventions on COVID-19 deaths (baseline model). A) Relative deaths difference (RDD) for vaccines. B) RDD for NPIs. The boxplots in both panels show the results considering 1000 stochastic trajectories in each region. The horizontal line within each box marks the median value, while the top and bottom edges correspond to the 90% CI. The whiskers extend to the maximum and minimum values. These estimates are obtained considering the baseline model.

Analogously, we compute relative difference of infections (RDI), defined as the fraction of total infections avoided by vaccines with respect to the infections observed in an equivalent model without vaccines (see section 3.3 in the Supplementary Information for details). In general, the RDIs are lower compared to RDDs across the regions as COVID-19 vaccines are more effective in preventing severe outcomes rather than infections. The RDIs show a similar pattern to that observed for RDDs across the four regions, except for São Paulo. The median of RDIs is only 9% while it is 51% for RDDs. This is likely due to São Paulo experiencing distinct viral strains, specifically the Gamma and Delta variants, which significantly reduces vaccine efficacy against infection compared to the other regions that instead saw the circulation of the wild type and the Alpha variant [44].

We also investigate when the vaccination starts to have macroscopic effects by estimating the time when weekly deaths with and without vaccination differ by more than 1%. The results show that the difference of deaths began to diverge after 18 weeks since the vaccine rollout in British Columbia, 6 weeks in Lombardy, 8 weeks in both London and São Paulo (see section 3.1 in the Supplementary Information). To investigate the effects of NPIs on the progression of the Pandemic, we run a counterfactual scenario where we remove the impact of NPIs on contacts, while maintaining vaccinations. By comparing the trajectory of deaths with/without contacts modulation induced by NPIs, we found that without NPIs we would have experienced a larger number of deaths across all regions considered. Specifically, our results show that removing NPIs would have resulted in a much higher peak of weekly deaths during the period considered, 5.3 (90% CI: [3.3, 9.4]) times higher in British Columbia, 8.8 (90% CI: [6.8, 11.9])times higher in Lombardy, 6.7 (90% CI: [5.5, 8.2]) times higher in London, and 4.7 (90% CI: [3.9, 5.7]) times higher in São Paulo compared to the estimates of the model considering NPIs. The absence of NPIs would have led to a 3 weeks earlier peak of weekly deaths in London and Lombardy, and 5 weeks São Paulo (see section 3.2 in the Supplementary Information). Furthermore, we quantify the effect of NPIs by computing the fraction of total deaths avoided by NPIs with respect to the deaths observed in an equivalent simulations without NPIs (denoted by RDD as above). As shown in Fig 2-B we find that 87.50% (90% CI:[82.82%,91.27%]) deaths have been avoided due to NPIs in British Columbia, 66.72% ([62.75%, 71.25%]) in Lombardy, 70.29% ([65.06%, 76.73%]) in London, and 35.07% ([28.82%, 45.12%]) in São Paulo. Not surprisingly, the RDD of the four regions are strongly correlated with their contact reduction with an exception in British Columbia. Specifically, British Columbia shows the highest RDD despite not featuring the strongest reduction in contacts. As noted above, the peak height of weekly deaths in British Columbia is more than five time smaller than the other regions. Hence, the NPIs in place, and their adoption, successfully managed to reduce the burden of the disease more than in the other locations. Thus, relatively speaking, without NPIs the picture in British Columbia would have been drastically different. The RDD of London is the second largest due to the strict NPIs implemented during the observed period, imposed by the emergence and spreading of the Alpha VOC in September 2020. In contrast, São Paulo exhibits the lowest RDD, due to the relative low reduction in contacts induced by NPIs.

Analogously, we compute the fraction of total infections avoided by NPIs with respect to the infections estimated by an equivalent model without NPIs (denoted by RDI as mentioned above, see details in section 3.3 in the Supplementary Information). The results of RDIs of the four regions are consistent with the results of RDD, except for London where we find slightly lower RDIs than Lombardy.

Comparing panels A and B in Fig. 2, we see that, in the first months of the vaccine rollout, NPIs averted more deaths than vaccinations underscoring the crucial role of NPIs in supporting the initial critical phases of mass vaccine campaigns that, as discussed, struggled with significant challenges in the first periods. Furthermore, the comparison between the two highlights the potential negative effects of behavioural relaxation mechanisms.

### Estimating the extent of behavioural relaxation induced by vaccines

Building on the baseline and the literature, we developed four behavioural models (<u>models 1-4</u>) where we incorporate behavioural relaxation mechanisms potentially induced by vaccines. In Fig. 3, we show the calibrated results of all models (including the baseline) by presenting the medians and the 90% confidence intervals of weekly deaths. The calibrated curves are all consistent with reported epidemiological data. The differences between the models appear minimal to a visual inspection. To better investigate the nuances, we computed weighted mean absolute percentage errors (wMAPEs) as well as the Akaike Information Criterion (AIC) [57], and the Bayesian Information Criterion (BIC) [32]. The wMAPE measures the difference between the median outcomes of our models and reported data, while AIC and BIC scores assess the performance of models by trading off their complexity and the goodness of fit. Based on the AIC/BIC scores, we further calculate the AIC/BIC weight of each model for a more intuitive interpretation [31, 32]. These weights can be interpreted as the probability that a model, among those considered, is the most likely given the empirical data [31, 32]. While we only display AIC/BIC weights in the main text, we refer the reader to section 5.1 in the Supplementary Information for the AIC/BIC scores.

**Fig 3.**
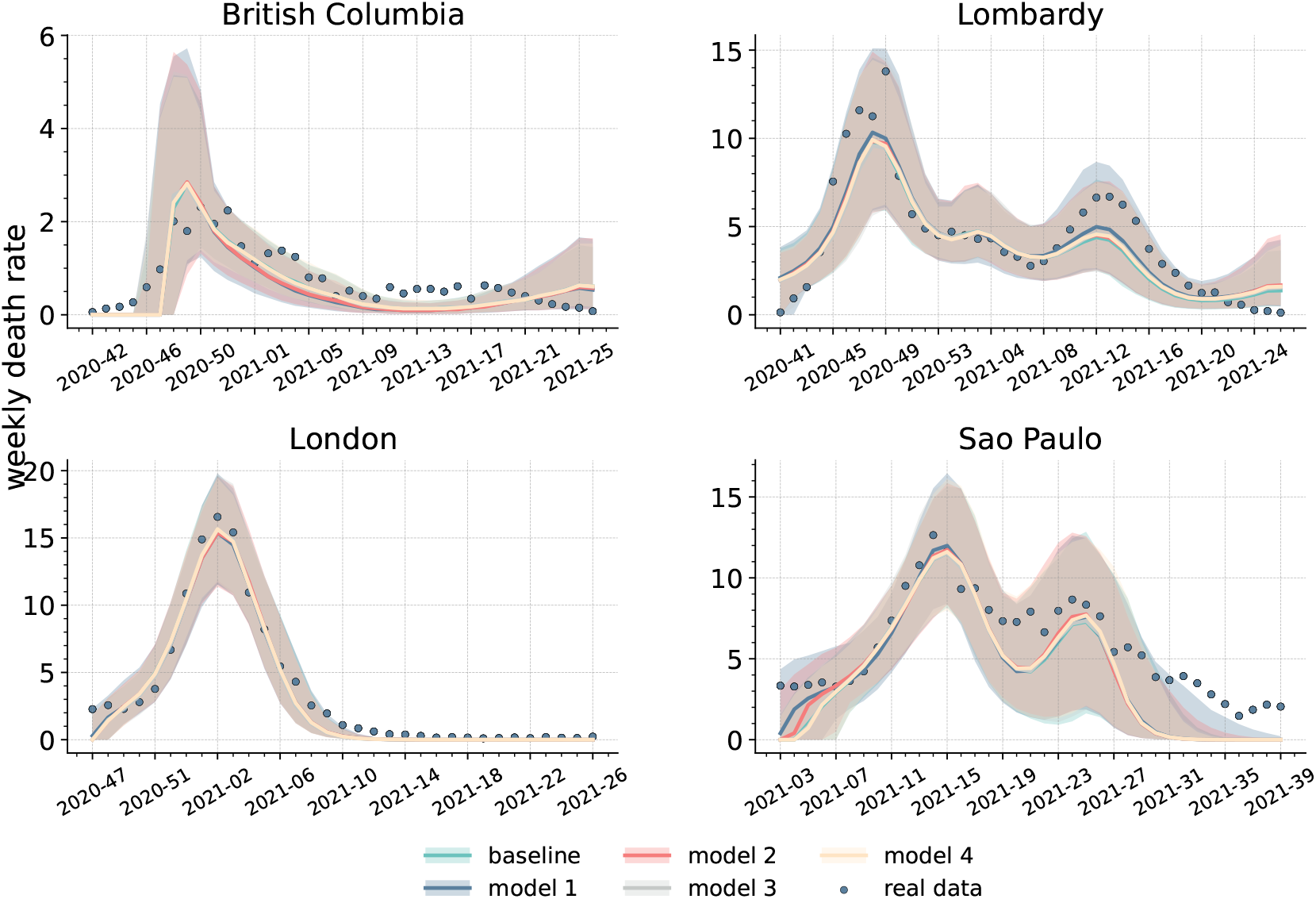
Comparison of behavioural and baseline models. Calibrated weekly deaths trajectories (i.e., weekly deaths per 100, 000) for the baseline and four behavioural models across the four regions. Solid lines indicate the medians, while the shaded areas the 90% confidence intervals. Reported weekly deaths are denoted by blue dots.

The wMAPEs of the median of the five models are shown in Table 1. Behavioural models lead to smaller errors than the baseline in three regions out of four, nonetheless the best model results in limited improvements. Indeed we find a decrease in wMAPE of 9.8% in Lombardy, 2.0% in London, and 6.1% in São Paulo compared to the baseline. In more detail, model 1, model 2, and model 3 achieve, respectively, the lowest wMAPE in Lombardy, São Paolo, and London.

**Table 1.** wMAPEs obtained comparing the medians of calibrated models and reported weekly deaths. The lowest wMAPE in each location, indicating best performance, is highlighted in bold.

| wMAMPE | baseline | model 1 | model 2 | model 3 | model 4 |
| --- | --- | --- | --- | --- | --- |
| British Columbia | **0.3751** | 0.4431 | 0.4406 | 0.3862 | 0.3833 |
| Lombardy | 0.2742 | **0.2473** | 0.2702 | 0.2760 | 0.2686 |
| London | 0.1508 | 0.1529 | 0.1561 | **0.1479** | 0.1483 |
| São Paulo | 0.3056 | 0.2889 | **0.2869** | 0.3009 | 0.2959 |

The picture changes, when we account for the complexity of the models. Indeed, according to AIC weights, the baseline is the most likely model in three regions (see Table 2). In Lombardy, instead, model 1 (followed by the baseline) emerges as the most likely. In the same table we can see that when considering BIC weights, the baseline is the most likely model across all regions. This metric is known to be more biased towards simpler model than the AIC. Nevertheless, in Lombardy the baseline is only marginally better than model 1. These results show that, although behavioural mechanisms improve the goodness of fit in three regions, they come at the cost of an increased complexity that does not always offset the gains of fits. Furthermore, due to the similarity of the outcomes, the selection of the most likely model is function of the metric considered (i.e., AIC vs BIC). Following Occam’s razor principle, we can conclude that the inclusion of behavioural relaxation mechanisms is not fully justified, at least when looking at weekly deaths in the four regions studied. A simpler model, that does not explicitly account for this phenomenon, appears well suited to reproduce the unfolding of reported deaths. As shown in section 5.2 of the Supplementary Information, the ranking of the models in terms of Akaike weights does not change by removing the last 1, 2, 3, 4 week(s). In the case of BIC weights, when removing the last 2, 3, 4 weeks we obtain model 1 to be more likely than the baseline, although the difference between the two remains small. In other words, the results are robust to the choice of the time horizon considered and are not affected by possible fluctuations in the tails of the epidemic curves.

**Table 2.** AIC and BIC weights computed considering calibrated models’ medians and reported weekly deaths. The highest AIC/BIC weight in each location is highlighted in bold.

| AIC | baseline | model 1 | model 2 | model 3 | model 4 |
| --- | --- | --- | --- | --- | --- |
| British Columbia | **0.95** | 0.00 | 0.00 | 0.03 | 0.02 |
| Lombardy | 0.09 | **0.88** | 0.02 | 0.00 | 0.01 |
| London | **0.84** | 0.02 | 0.02 | 0.04 | 0.08 |
| São Paulo | **0.64** | 0.12 | 0.13 | 0.05 | 0.06 |
| BIC |  |  |  |  |  |
| British Columbia | **0.99** | 0.00 | 0.00 | 0.00 | 0.00 |
| Lombardy | **0.54** | 0.44 | 0.01 | 0.00 | 0.01 |
| London | **0.98** | 0.00 | 0.00 | 0.01 | 0.01 |
| Sao Paulo | **0.95** | 0.02 | 0.02 | 0.01 | 0.01 |

**Table 3.** Relative deaths difference for behavioural mechanisms. Medians and 50% confidence intervals are reported. Numbers indicate percentages.

| RDD (%) | model 1 | model 2 | model 3 | model 4 |
| --- | --- | --- | --- | --- |
| British Columbia | -0.48 [-4.73,3.54] | -0.12 [-3.26,2.58] | 0.27 [-2.59,3.02] | 0.10 [-1.83,1.92] |
| Lombardy | -4.34 [-16.46,-0.79] | -0.58 [-1.67,0.22] | -0.33 [-1.08,0.31] | -0.14 [-0.77,0.39] |
| London | -1.91 [-8.57,-0.34] | -0.21 [-0.85,0.29] | -0.01 [-0.53,0.52] | 0.00 [-0.23,0.24] |
| São Paulo | -30.35 [-67.68,-7.91] | -5.18 [-25.42,-0.23] | -1.01 [-2.69,-0.2] | -0.45 [-1.5,0.03] |

**Table 4.** Model parameters.

| Parameter | Symbol | Value |
| --- | --- | --- |
| Reproductive number | $R_0$ | Calibrated within $[1, 3]$. |
| Latent period | $\epsilon$ | $3^{-1}days^{-1}$ for Delta [52],<br>$3.7^{-1}days^{-1}$ for others[48–51] |
| Infectious period | $\mu$ | $2.5^{-1}days^{-1}$ [48, 70] |
| Transmission rate | $\beta$ | Obtained from $R_0$ |
| Infection fatality rate | $IFR$ | Ref.[58] |
| Contact matrix | $\mathbf{C}$ | Ref. [65] |
| Delayed days in reporting deaths | $\Delta$ | Calibrated within $[3, 64]$ [69] |
| Initial fraction of infections | $i_{ini}$ | Calibrated within $[0.0005, 0.02]$ |
| Initial fraction of recoveries | $r_{ini}$ | Calibrated within $[0.1, 0.4]$ |
| Adjustment of the start date of the simulation of epidemic with respect to the baseline date ($t^*$) | $\Delta t$ | Calibrated within $[0, 7]$ weeks |
| Adjustment of the introduction date of a VOC with respect to the date ($t_{var}^*$) at which each variant is consistently featured in the genomics data | $\Delta t_{var}$ | Calibrated within $[0, 42]$ days [71] |
| Relative transmissibility of a second variant | $\sigma$ | 1.5 for Alpha [44];<br>calibrated within $[1.0 - 2.5]$ for Delta with respect to Gamma [43] |
| Overall vaccine efficacy | $VE$ | 0.9 against wild type and Delta variant;<br>0.85 against Alpha variant;<br>0.8 against Gamma variant [46, 47] |
| Vaccine efficacy against infection | $VE_S$ | 0.85 against wild type;<br>0.75 against Alpha variant;<br>0.65 against Gamma variant;<br>0.6 against Delta variant[46, 47] |
| Transition rate towards non-compliance | $\alpha$ | Calibrated within $[0.0001 - 10]$ |
| Transition rate towards compliance | $\gamma$ | Calibrated within $[0.0001 - 10]$ |
| Relative infection risk of non-compliant individuals | $r$ | Calibrated within $[1.0 - 1.5]$ |

As shown in section 5.4 in Supplementary Information, the posterior distributions of increased transmissibility (i.e., *r*) of non-compliant individuals range between 10 − 30%, which is a significant increase. However, with the exception of São Paulo in case of model 1, the median fraction of non-compliant individuals at each time step is firmly smaller than 20% (see section 4.2 in Supplementary Information). In other words, the calibration selects regions of the phase space where the population of non-compliant individuals is the clear minority. This result corroborates the lack of clear signs of behavioural relaxation on weekly deaths. Indeed, even assuming the presence of such phenomenon, the empirical evidence constraints it to a small group of the total population.

In order to gather a better understanding of the dynamics at play, and to isolate the potential effects of behavioural relaxation on deaths, we run another counterfactual analysis removing the relaxation mechanisms in the four calibrated behavioural models. In doing so, we compute the relative deaths difference (RDD) between the models with/without behavioural relaxation. As shown in Fig. 4 and in Table 5, the median RDD values are below zero in the large majority of cases, indicating that removing behavioural relaxation generally results in fewer deaths. The results show that models 1 and 2 lead to a larger difference in deaths (especially model 1 in São Paulo) compared to models 3 and 4 which restrict behavioural relaxation to vaccinated individuals only. Not surprisingly, the impact of non-compliance extended to the whole population is higher. It is important to notice how, with two exceptions (i.e., model 1 and model 2 in São Paulo), the RDD values are close to zero. As noted above, the posterior distributions of behavioural parameters selected in the calibration lead to configurations where relaxation does not strongly impact deaths.

**Fig 4.**
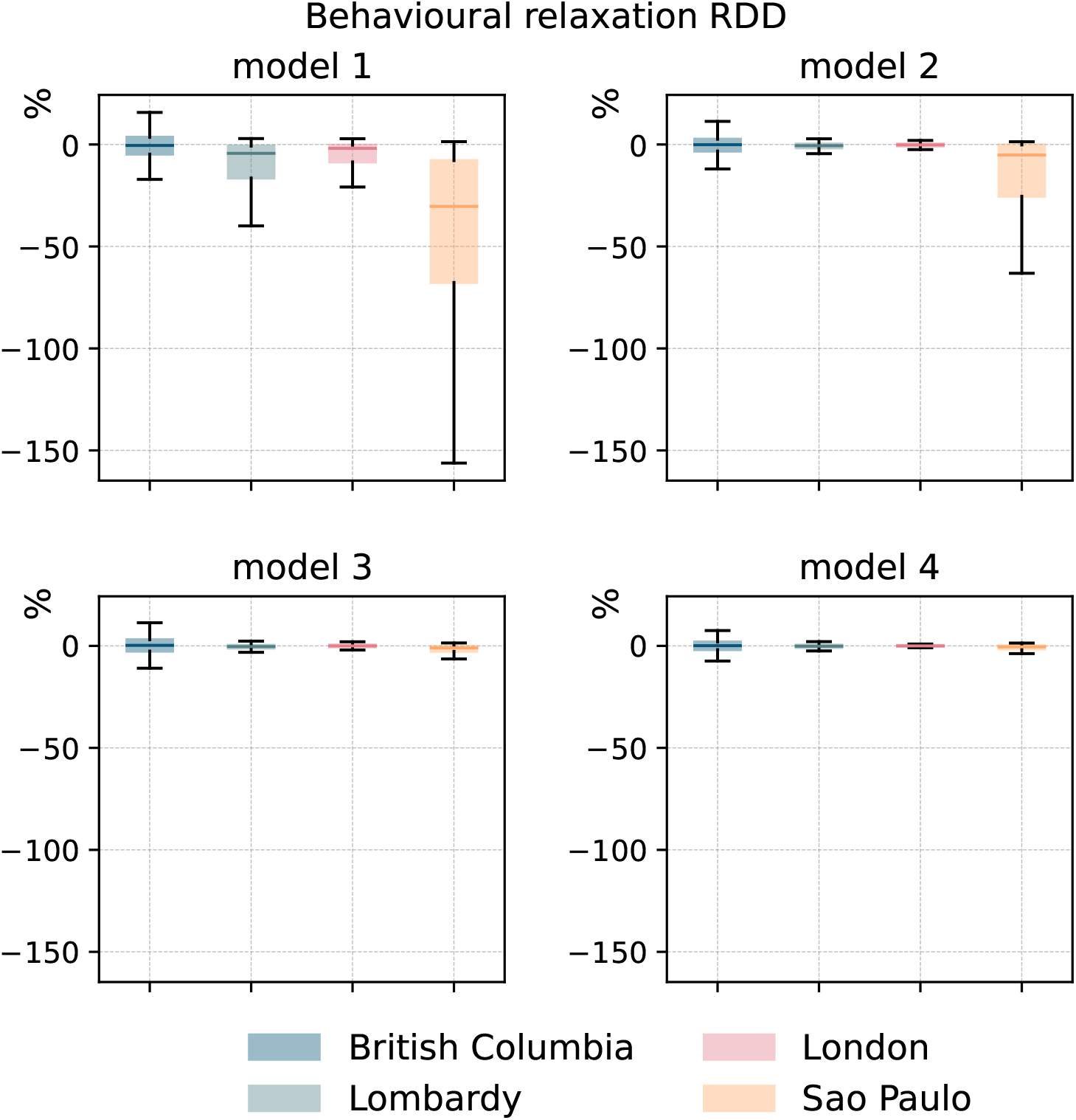
The impact of behavioural relaxation on COVID-19 deaths (behavioural models). We plot the RDD (i.e., relative deaths difference) for behavioural mechanisms in the four models and regions. Each boxplot is built considering 1000 stochastic simulations. The horizontal line within each box marks the median value, while the top and bottom edges correspond to the 50% CI. The whiskers extend to the maximum and minimum values after removing the outliers that beyond 1.5 times the interquartile range.

**Table 5.** Relative infection difference in counterfactual scenarios without behavioural relaxation.

| RDI (%) | model 1 | model 2 | model 3 | model 4 |
| --- | --- | --- | --- | --- |
| British Columbia | -0.81 [-11.85,6.93] | -3.04 [-13.47,3.19] | -0.3 [-7.13,6.16] | -0.27 [-4.64,3.41] |
| Lombardy | -5.18 [-19.08,-0.86] | -0.78 [-2.96,0.0] | -0.52 [-1.38,0.04] | -0.27 [-0.91,0.21] |
| London | -1.39 [-7.36,-0.17] | -0.04 [-0.25,0.01] | -0.0 [-0.05,0.05] | -0.0 [-0.02,0.02] |
| Sao Paulo | -2.08 [-7.44,-0.12] | -1.54 [-7.76,-0.01] | -0.46 [-1.42,-0.08] | -0.15 [-0.83,-0.03] |

Finally, we compute the relative infection difference (RDI) (shown in section 3.4 of the Supplementary Information), which shows consistent results with those of RDDs.

## Discussion

In this paper, we aimed to find signs, and to quantify the extent of behavioural relaxation possibly induced by vaccines during the initial phase of the COVID-19 vaccine rollout. To this end, we developed a series of stochastic epidemic compartmental models integrating age-structure, vaccinations, NPIs, variants of concern, and deaths. We used a baseline model, without any behavioural relaxation mechanism, as a reference, and on top of this, we developed four behavioural models that extended previous work to account for individual behaviours in response to vaccination and the epidemic [21–26]. We tested these models considering weekly deaths in four regions: British Columbia (Canada), Lombardy (Italy), London (United Kingdom), and São Paulo (Brazil). These locations sample different epidemic, socioeconomic and socio-demographic contexts, as well as different vaccine rollout schedules and coverages. We first calibrated the baseline model to reported data and studied two counterfactual scenarios to quantify the impact of vaccines and NPIs on COVID-19 deaths and infections. Our results confirmed that both significantly reduced mortality and infections. Furthermore, they highlighted the critical role of NPIs in supporting the challenging initial phases of vaccinations. We then calibrated the four behavioural models and compared them considering both their goodness of fit and complexity. Behavioural models estimates are closer to real data than the baseline in three locations, though the improvements are limited between 2% and 10% in terms of wMAPE. Behavioural mechanisms increase models’ complexity which is not always offset by the benefits of improved fits. This suggests that additional mechanisms of behavioural relaxation linked to vaccination may not be evident across all regions. Furthermore, our results suggest that, even if behavioural relaxation took place, it was limited to a firm minority of the population.

Overall, our results indicate that behavioural relaxation did not leave clear marks on reported deaths. This finding, in line with surveys conducted in France [30], might be interpreted as a lack of support for systematic behavioural relaxation induced by COVID-19 vaccines. However, our findings do not exclude that, in fact, behavioural relaxation took place as suggested by other surveys [27–29]. As mentioned above, assuming the presence of behavioural relaxation, the calibration with weekly deaths constraints the phase space to regions where the fraction of non-compliant individuals is a firm minority. This might explain the good performance of the baseline: the impact of behavioural relaxation could be accounted for by simpler models that do not explicitly consider additional mechanisms. In our settings, the effects of behavioural relaxation might be fully captured by the modulation of contacts induced by NPIs.

It is important to mention how the selected target variable (i.e., weekly deaths) might have influenced our findings. Indeed, behavioural relaxation might have been more prevalent but not to the levels needed to affect mortality levels observed at a macroscopic scale. Signs of behavioural relaxation might be clearer in other indicators. For example, given the strong dependence of COVID-19 mortality on age [58], behavioural relaxation might have primarily affected infections, especially among the young, active population, rather than deaths. However, data on confirmed cases has been shown to be a poor indicator and a very hard signal to fit due to under-reporting and variations in testing policies among other factors [59].

These possible interpretations of our results highlight the issue of non-identifiability of complex behavioural mechanisms in epidemic models linked to highly degenerate phase-spaces and to the interplay among the various processes at hand (e.g., disease transmission and behavioural reactions) [60]. Arguably, the quest towards a clear identification of behavioural reactions to epidemics is linked to the use of multi-stage calibration steps informed by a range of data types and indicators that go beyond the solely use of epidemic variables [60]. Progresses in this direction are contingent to advances in data collection and data sharing as well as to the identification of key behavioural observables and novel data streams to track [61, 62].

Our work comes with limitations. First, the epidemiological and vaccination data are sourced from different datasets. Although the data has been obtained from official sources, the granularity provided is not homogeneous. Second, we considered a simplified vaccination protocol assuming that only susceptible individuals can get vaccinated and a single dose regiment. These assumptions have been made to simplify the model structure. Third, we used regional-level data regarding school closures for British Columbia, London, and São Paulo but country-level data for Lombardy, due to the lack of specific regional data within Italy. Moreover, there is lack of available data to parametrize the rates regulating behavioural relaxation. As a consequence, we had to calibrate the behavioural parameters within some rather arbitrary ranges. Fourth, though we accounted for a higher transmission rate, shorter latent period, and decreased vaccine efficacy for the second variant, we used the same infection fatality rate (IFR) across all strains and regions. Finally, our model does not account for socioeconomic nor socio-demographic differences in vaccines uptake nor in adoption of NPIs [63, 64].

Overall, our work highlights the critical importance of both NPIs and vaccines in curbing COVID-19 deaths and infections during the initial months of COVID-19 vaccination campaigns. Our findings pave the way for further research to refine the proposed models and deepen our understanding of the interaction between individual protective behaviour and vaccinations in a broader context.

## Materials and Methods

### Baseline model

As baseline we adopt a stochastic age-stratified epidemic compartmental model that integrates vaccination, NPIs, and the emergence/spread of a second variant based on a Susceptible-Latent-Infected-Recovered (SLIR) compartmentalization extended to account for deaths. Individuals are grouped into 16 age brackets with a five-year interval (except for the last group which is 75+). We use age-stratified Infection Fatality rates (IFR) from Ref. [58] and age-stratified contact matrices from Ref. [65]. The natural history of the disease is modelled as follow. By interacting with the Infected (*I*), Susceptible (*S*) individuals transition to the latent stage (*L* compartment) where they are infected but not yet infectious. We assume a force of infection (i.e., the rate at which Susceptible get infected) as a function of age, transmissibility of each strain, contact matrices, and NPIs (see below for details). Individuals stay in *L* for an average of *ϵ*^−1^ days. After, they become infectious transitioning to the *I* compartment. After the infectious period *μ*^−1^, infected individuals either recover with probability (1 − *IFR*_*k*_) (transitioning to the *R* compartment) or die from the disease with probability *IFR*_*k*_ (transitioning to the *D* compartment), where *k* denotes the age-group. We also consider a delay of Δ days in reporting deaths. Therefore, individuals are moved to the *D*^*o*^ compartment from *D* after Δ days.

### Modelling vaccinations

We incorporate vaccinations into our models by doubling all the compartments to include vaccinated individuals at any stage of the disease. We assume that only susceptible individuals can receive the vaccine. Additionally, to simplify the model, we disregard the time interval between the first and second dose. Consequently, we assume that individuals acquire full protection right after the first inoculation. We use real data to capture the unfolding of the vaccination campaigns in the four regions [34–38]. Notably, vaccine data are recorded weekly in official statistics in British Columbia while the other three regions report vaccine data daily. Thus, in British Columbia, we convert the weekly doses into daily by splitting them homogeneously in each week day. Vaccines protect individuals in two ways: by lowering the risk of infection, and by reducing the risk of death in case of breakthrough infection. In practice, for vaccinated individuals the force of infection is multiplied by a factor 1 − *V E*_*S*_, where *V E*_*S*_ denotes the vaccine’s efficacy against infection. If a vaccinated individual becomes infected, the IFR is further reduced by a factor 1 − *V E*_*M*_, where *V E*_*M*_ represents the vaccine’s efficacy against death. Therefore, the overall vaccine efficacy for a susceptible individual, against death, is *V E* = 1 − (1 − *V E*_*S*_)(1 − *V E*_*M*_).

### Modelling the impact of NPIs on contacts

In age-structured epidemic models, contact matrices **C** stratify interactions among age groups [65]. Contact matrices might be further stratified by the context (i.e., location) where contacts take place: home, school, workplace, and general community settings [66]. Here, we consider both dimensions and express the overall contact matrix as the sum of the contact matrices of each context:

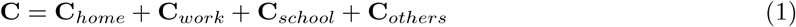

To estimate the variations in contact rates due to NPIs, we use mobility data from the COVID-19 Community Mobility Report released by Google [39] and the Oxford Coronavirus Government Response Tracker (OxCGRT) [40]. In particular, we use this data to adjust the contact matrix as follows:

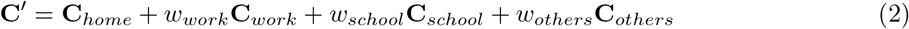

where **C**^′^ is the adjusted contact matrix, *w*_*work*_ and *w*_*others*_ are computed as (1 − *x/*100)^2^, where *x* is the percentage of change in mobility with respect to a pre-Pandemic baseline measured by Google [39]. We use a square form as the number of contacts in a location scales proportional to the square of people visiting that location [5, 6]. In particular, *w*_*work*_ is computed considering the field related to workplaces of the Google Mobility Report, while *w*_*others*_ considering an average of the fields related to retail and recreation and transit stations. Furthermore, *w*_*school*_ is computed as (3 − *school*)/3, where *school* is an index measuring the strictness of containment policies in schools computed by OxCGRT [40]. It takes integer values from 0 (i.e., no containment measures are in place) to 3 (i.e, full school closure). Contacts at home are not adjusted although we acknowledge they may increase due to the adoption of NPIs. The factor representing the contact level with respect to the pre-pandemic level in Fig 1 is computed as (*w*_*work*_ + *w*_*school*_ + *w*_*others*_)/3.

### Modelling multiple viral strains

In order to capture the spread of a second variant in British Columbia, Lombardy, and São Paulo, we introduce new compartments *L*^′^, *I*^′^, *R*^′^, *D*^′^ and 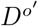 for non-vaccinated individuals infected by a second strain. Similarly, we consider compartments 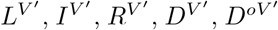 for vaccinated individuals infected by a VOC. We model the introduction of a second variant as follows. We denote *t*_*var*_ as the time at which the second variant establishes its presence in each location. To this end, we use genomics data from Ref. [41] and calibrate *t*_*var*_ in a range between 0 and 42 days prior to the first date in which each variant is consistently featured in the genomics data (i.e., the share of samples attributed to the variant in the genomics data is greater zero from this first date onwards). We did not consider the first appearance in the dataset as a single sample might be linked to an isolated importation from a different location. Then, during simulations at *t*_*var*_, we initialize the compartment *I*^′^ by allocating there 1% of infected individuals (considering both *I* and *I*_*V*_).

The four regions studied faced the Alpha, Gamma, and Delta VOC. In detail, in British Columbia and Lombardy, Alpha appeared as a second strain replacing the wild type. In London, the Alpha variant was the dominant variant circulating throughout our time horizon. In São Paulo, the initial variant observed at the start of the simulation period was Gamma which was then replaced by Delta.

We model VOCs by adjusting (or not) relevant parameters. According to literature, the latent period of Alpha and Gamma is similar to that of the wild type [48–51]. Hence, we kept *ϵ*^′^ = 3.7^−1^*days*^−1^ for these variants. In contrast, the latent period of Delta has been reported to be shorter [52]. Hence, we set *ϵ*^′^ = 3^−1^*days*^−1^ for this VOC. For all variants, including the wild type, we set the infectious period to *μ* = 2.5^−1^*days*^−1^. Additionally, the second variant may exhibit higher transmissibility. Thus we adjust the transmissibility of variants by multiplying it by a parameter *σ*, which represents the relative increase in transmissibility. Following the literature we set *σ* = 1.5 for Alpha compared to the wild type [44]. In São Paulo, where Delta replaced Gamma, we calibrate *σ* in a range of [1.6 − 2.5], as no specific indication was found in the literature. Moreover, the vaccine efficacy might also be lower against variants [44, 46, 47]. Following the literature, we set the vaccine efficacy as *V E* = 90% (*V E*_*S*_=85%) against the wild type [46], *V E* = 85% (*V E*_*S*_ = 75%) against Alpha [46], *V E* = 80% (*V E*_*S*_ = 65%) against Gamma [47], and *V E* = 90% (*V E*_*S*_ = 60%) against Delta [46].

### Behavioural models

Building on the baseline and the literature, we implemented four additional models that also include behavioural relaxation mechanisms [27, 28]. To this end, we introduce new compartments 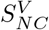 and *S*_*NC*_ to account for susceptible individuals (vaccinated or not) that relax adoption of NPIs becoming non-compliant (NC). In detail, individuals who relax their behaviour transit from *S* (*S*^*V*^) to 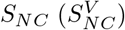. Conversely, non-compliant individuals who return to compliant behaviours transition from 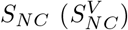 to *S* (*S*^*V*^). We assume that individuals in the non-compliant compartments get infected at higher rates compared to those in compliant compartments [23, 24]. This is accounted for by multiplying their force of infection by a factor *r* > 1.

Following the literature, we study four different behavioural models that differ in the mechanisms used to describe how individuals enter and leave the non-compliant compartments. In <u>model 1</u>, susceptible individuals (vaccinated or not) can enter or leave the NC compartments at constant rates *α* and *γ*. In <u>model 2</u>, susceptible individuals enter or leave the NC compartment at varying rates. The transition rate from *S* (*S*^*V*^) to 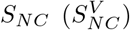 is set as a function of the fraction of vaccinated individuals and a parameter *α*. The transition rate from 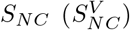 to *S* (*S*^*V*^) is set as a function of the number of reported daily deaths per 100, 000 and a parameter *γ*. <u>Model 3</u> and <u>model 4</u> are analogous to model 1 and 2. However, only vaccinated individuals can transition to the NC compartment. The structure of our models is illustrated in Fig. 5. More details are reported in the Supporting Information.

**Fig 5.**
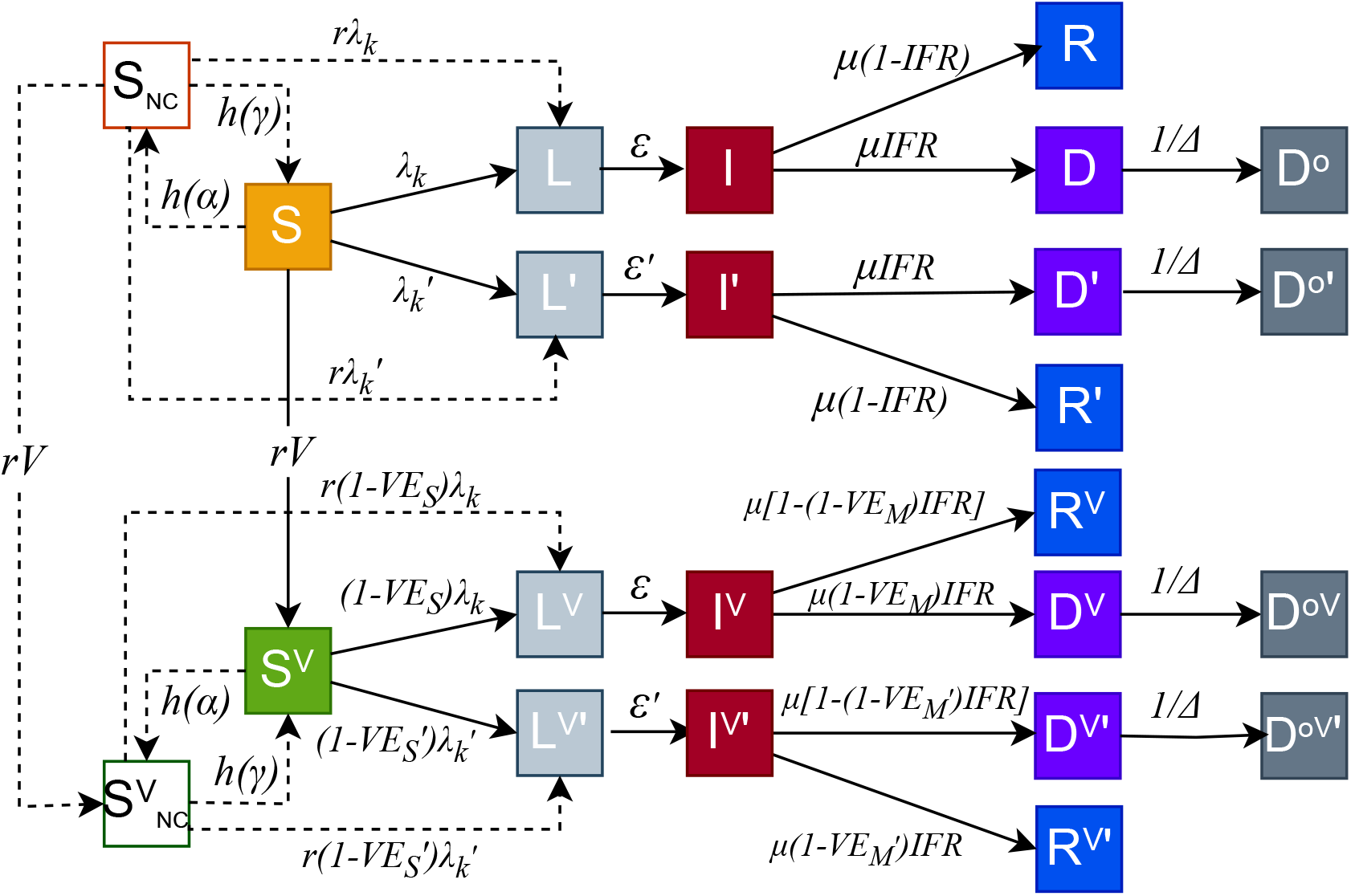
Epidemic compartment structure. All compartments connected by solid lines constitutes the baseline model. This model includes susceptible (*S*), latent (*L*), infected (*I*), recovered (*R*), and dead (*D, D*^*o*^) compartments. The top row represents non-vaccinated compartments, whereas the bottom row represents vaccinated compartments. Individuals in the *S* compartment get vaccinated according to real vaccine rates (*rV*) and then transition to the *S*^*V*^ compartment. To account for the emergence of a second variant, we double the compartments creating *L*^′^, *I*^′^, *R*^′^, *D*^′^, and 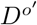. This is done also for the vaccinated compartments that become 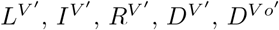. Behavioural models include susceptible non-compliant compartments (*S*_*NC*_, 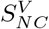) connected by dotted lines, where individuals have *r* times higher probability of getting infected with respect to susceptible compliant individuals (S and *S*^*V*^). In models 1 and 2, we include 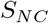 and 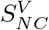, whereas in models 3 and 4 we include only 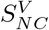.

### Models calibration

We apply an Approximate Bayesian Computation-Sequential Monte Carlo (ABC-SMC) method to calibrate our models [33] to reported data. The goal of ABC-SMC algorithm is to estimate the posterior distribution of free parameters ***θ*** starting from an input prior distribution *P*(***θ***). It is an extension of the ABC rejection algorithm, where suitable parameters are found by iteratively sampling from the prior distribution and computing for each sampled parameter set ***θ***_*i*_ a distance function *d*(**y**_*i*_, **y**_*data*_), where **y**_*i*_ ∼ *f* (*θ*_*i*_) is the output of the model and **y**_*data*_ is the reported data (i.e., weekly deaths). Each *θ*_*i*_ is accepted if *d*(**y**_*i*_, **y**_*data*_) ≤ *ξ*, where *ξ* is a predefined tolerance. The process is repeated until *M* parameters *θ*_*i*_ are accepted. Their distribution approximates the true posterior distribution Π(*θ*|**y**_*data*_, *ξ*). The ABC rejection algorithm is of straightforward implementation, however suffers from several limitations. First, the values of *M* and of the tolerance *ξ* are free parameters that shape the interplay between convergence speed and accuracy [67]. Second, the prior distribution is never updated to account for information from previous iterations. The ABC-SMC framework has been developed to tackle these issues. It consists of *T* generations (i.e., iterations). The first one is based on a rejection algorithm step where *ξ* is set to a high value. In the second generation, the tolerance is decreased, parameters are sampled from those accepted in the previous step and perturbed via a kernel to avoid converging on local minima of the phase space. The process is repeated for *T* generations of *M* particles (i.e, samples) each. Then, the set of accepted parameters in the last generation is used as the empirical posterior distribution. We adopted a python implementation of ABC-SMC from the library <u>pyabc</u> [68].

The free parameters and the priors explored in our models are:

- Reproductive number *R*_0_. We explore values in the interval [1, 3].
- Delay in reporting deaths Δ. Consistent with observations, we explore the interval [3, 64] [69].
- Initial fraction of infections of the total population *i*_*ini*_. We estimating the ranges from the number of deaths and IFR across the four regions. We explore the interval [0.0005, 0.02].
- Initial fraction of individuals with residual immunity from past waves *r*_*ini*_. We explore the range of [0.1, 0.4].
- Start date *t*_0_ of the simulation of epidemic. We calibrate *t*_0_ within a range of 8 weeks such that *t*_0_ = *t*^∗^ − Δ*t*, where Δ*t* = [0, 1, …, 7] week(s). The baseline dates *t*^∗^ are set as 8 weeks before the peak of mortality in real data. Following this, *t*^∗^ is set as 2020/10/12 for British Columbia, 2020/10/05 for Lombardy, 2020/11/16 for London, and 2021/01/18 for São Paulo.
- Introduction date of a VOC *t*_*var*_ (applied for all regions except London). we use genomics data from Ref. [41] and calibrate *t*_*var*_ in a range between 0 and 42 days prior to the first date 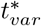 from which each variant is consistently featured in the genomics data. Thus, 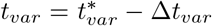, where Δ*t*_*var*_ = [0, 1, …, 42] day(s). 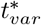 is set as 2020/12/21 in British Columbia, 2020/9/28 in Lombardy, and 2021/3/29 in São Paulo.
- Relative transmissibility *σ* of Delta with respect to Gamma. In the case of São Paulo, Delta replaced Gamma. Literature shows that Delta is about 1.3 times more transmissible than Gamma [43]. We explore the interval [1.0 − 2.5].
- Behavioural parameters *α* and *γ*. We explore the interval [0.0001 − 10]. For both and sample them on a logarithmic scale.
- Relative infection probability of non-compliant individuals *r*. Individuals who relax their behaviour are more likely infected. Therefore, we increase the infection probability of non-compliant individuals by multiplying it by a factor *r*. We explore the interval [1.0, 1.5].

The initial prior distribution *P*(***θ***) is obtained sampling each interval uniformly.

### Model initialization

We initialize the number of individuals in each compartment as follows. We assume that, at the beginning, all individuals are in the compartments *S* or *L* or *I* or *R*. The initial individual numbers of infected (including both *L* and *I* compartments) and recovered (*R*) individuals are set as fractions of total population considering under-reporting and official data. The total number of infected individuals is then distributed to *L* and *I* compartments proportionally to the inverse of their respective transition rates. Besides, since our model is age-stratified, the initial numbers of individuals in compartments *S* (*L* or *I* or *R*) in each age group is set as *N*_*S*_ × *N*_*k*_*/N* (*N*_*L*_ × *N*_*k*_*/N* or *N*_*I*_ × *N*_*k*_*/N* or *N*_*R*_ × *N*_*k*_*/N*), where *N*_*k*_ is the individual number in age-group *k, N* is the total individual number, *N*_*S*_ is the total number of individuals in compartment *S*. All parameters of our models are displayed in Table 4.

### Models evaluation

We utilize weighted mean absolute percentage errors (wMAPEs), Akaike Information Criterion (AIC) scores [57], and Bayesian Information Criterion (BIC) scores [32] for evaluating the five models. wMAPE measures the difference between the median outcomes of our models and reported data. It is defined as:

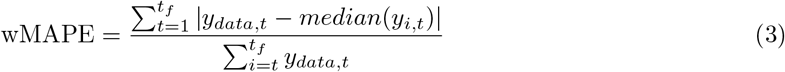

where *y*_*data*,*t*_ is the reported data at time *t, median*(*y*_*i*,*t*_) is median trajectory of model *i* at time *t*, and *t*_*f*_ is the total number of weeks.

AIC scores assess the performance by trading off the complexity and fitting of the models. The AIC score of model *i* is computed as:

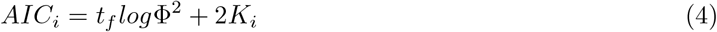

where Φ^2^ is the sum of the squares of residuals, *t*_*f*_ is the number of data points (i.e., weeks considered), and *K*_*i*_ is the number of free parameters of model *i*. To obtain a more intuitive metric, we calculate Akaike weights from the AIC scores. These can be interpreted as the relative likelihood of a given model [31]. The Akaike weight of model *i*, denoted by *w*_*i*_, is computed as

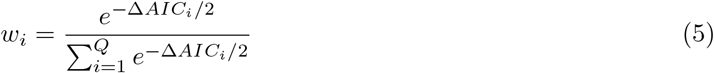

where Δ*AIC*_*i*_ is the difference between the AIC score of model *i* and of the best model (i.e., the one with lowest AIC score), and *Q* is the number of models.

BIC scores are similar to AIC scores, but contain a different term for measuring models’ complexity. The BIC score of model *i* is computed as:

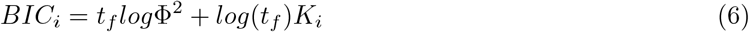

To obtain a more interpretable comparison of models, we also calculate BIC weights in an analogous way to Akaike weights. The BIC weight of model *i*, denoted by *w*_*i*_, is computed as

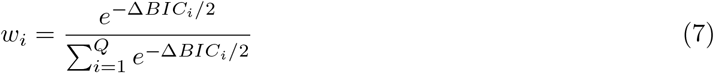

where Δ*BIC*_*i*_ is the difference between the BIC score of model *i* and of the best model (i.e., the one with lowest BIC score), and *Q* is the number of models.

### Relative deaths difference

To quantify the effect of vaccination, NPIs or behavioural relaxation on deaths, we compute the relative deaths difference (RDD) as the relative difference between the total number of deaths as simulated by the original model and in a counterfactual scenario where vaccinations, NPIs, or behavioural relaxation are removed. The relative deaths difference is calculated as:

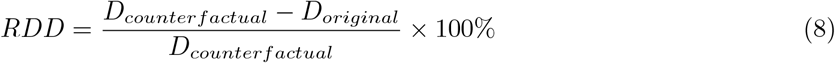

where *D*_*original*_ and *D*_*counterfactual*_ are the total number of deaths simulated in the original model and in the counterfactual scenario, respectively. The same approach, applied to infections, is used to compute the relative different of infections (RDI).

## Data Availability

All data produced are available online at https://github.com/Jadecoool/Estimating-behavioural-relaxation-induced-by-COVID-19-vaccines-in-the-first-months-of-their-rollout/tree/main

## Acknowledgments

Y.L. acknowledges support from the China Scholarship Council (CSC). N.G. acknowledges support from the Lagrange Project of the Institute for Scientific Interchange Foundation (ISI Foundation) funded by Fondazione Cassa di Risparmio di Torino (Fondazione CRT). All authors thank the High Performance Computing facilities at Queen Mary University of London.

## Author Contributions

Y.L., N.G, and N.P. designed the research. Y.L. performed the simulations. Y.L., and N.P. wrote the first draft of the manuscript. All authors contributed interpreting the data, editing and approving the manuscript.

## Data and code

The data and code to replicate the results can be found in https://github.com/Jadecoool/Estimating-behavioural-relaxation-induced-by-COVID-19-vaccines-in-the-first-months-of-their-rollout/tree/main

## Supporting information

### Epidemic models

All the epidemic models studied here are based and built on a baseline which is a stochastic age-stratified epidemic compartmental model that integrates vaccination, NPIs, and the emergence/spread of a second variant. We consider a Susceptible-Latent-Infected-Recovered (SLIR) compartmentalization with the addition of deaths. Individuals are grouped into 16 age brackets with a five-year interval (except for the last 75+ group). We use age-stratified Infection Fatality rates (IFR) from Ref. [58] and age-stratified contact matrices from Ref. [65]. The natural history of the disease is modelled as follow. Susceptible individuals (*S*) transition to the latent stage (*L* compartment) where they are infected but not yet infectious. We assume a force of infection (i.e., the rate at which *S* get infected) function of age, transmissibility of each strain, contact matrices, and NPIs (see below for details). Individuals stay in *L* for an average of *ϵ*^−1^ *days*^−1^. After, they become infectious thus transitioning to the *I* compartment. After the infectious period *μ*^−1^, individuals either recover with the probability (1 − *IFR*_*k*_) (transitioning to *R*) or die from the disease with probability *IFR*_*k*_ (transitioning to *D*), where *k* denotes the age-group. We also consider a delay of Δ days in reporting deaths. Therefore, individuals are moved to compartment *D*^*o*^ from *D* after Δ days, capturing the delay in deaths reporting.

We simulate the disease progression by using stochastic chain binomial processes in all models. For age group *k* the baseline model is defined by the following set of stochastic equations:

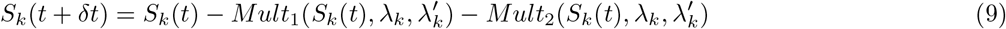

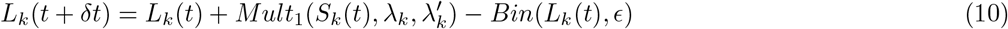

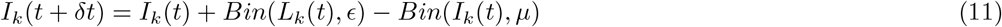

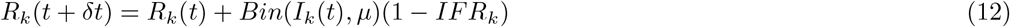

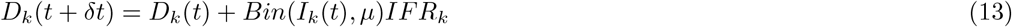

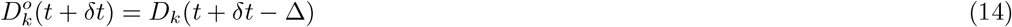

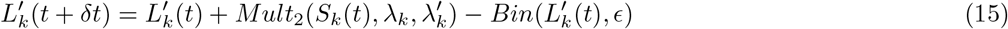

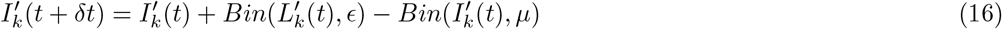

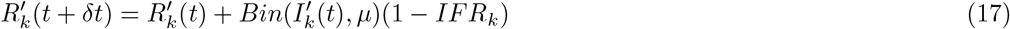

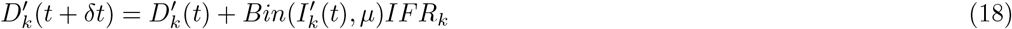

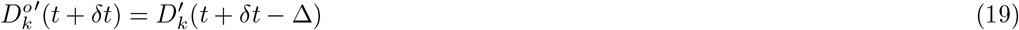

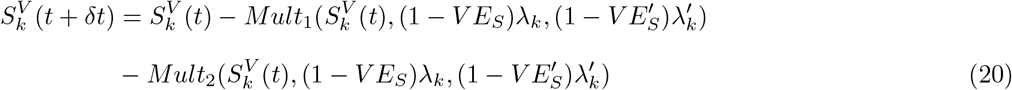

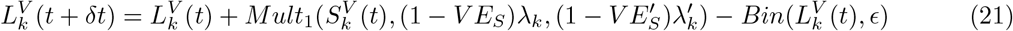

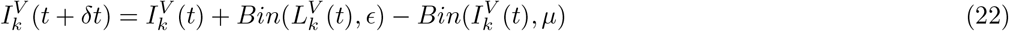

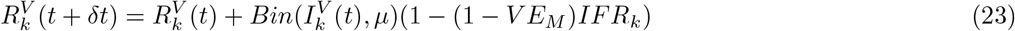

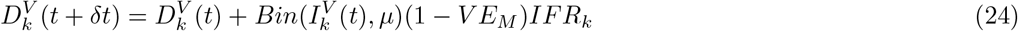

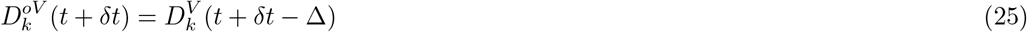

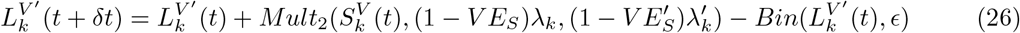

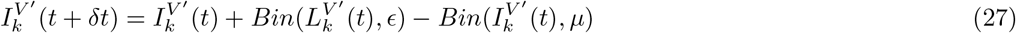

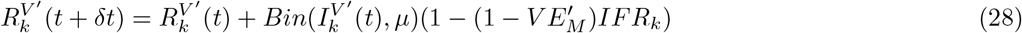

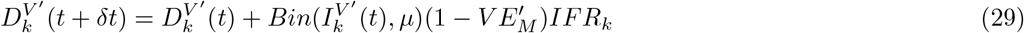

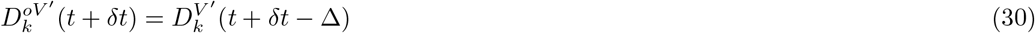

where the force of infection of the original strain is

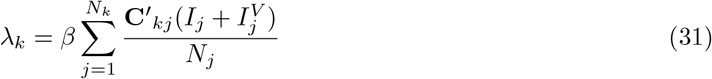

while is the force of infection of the second strain (if any) is

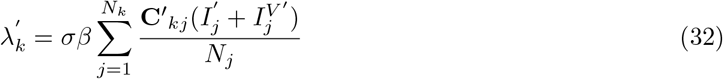

*β* is the transmission rate of the first strain, *σ* indicates the relative transmissibility of the second strain compared to the previously circulating one. **C**^′^ is the contact matrix adjusted for NPIs. To avoid issues with transition probabilities large than one, we transform the rates λ_*k*_ and 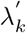 with the function *f* (λ) = 1 − *e*^−λ^. Furthermore, *Mult*_1_(*X, p*_1_, *p*_2_) and *Mult*_2_(*X, p*_1_, *p*_2_) describe, respectively, a draw from the random variable 1 occurring with probability *p*_1_ and the random variable 2 occurring with probability *p*_2_, given *X* trials.

In the behavioural models we add non-compliant compartments. In models 1 and 2 non vaccinated and vaccinated susceptible can both become non-compliant (transitioning to, respectively, *S*_*NC*_ and 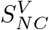). In models 3 and 4 the behavioural relaxation is linked only to vaccinated susceptible individuals.

The transitions among compartments in models 1 and 2 are as following:

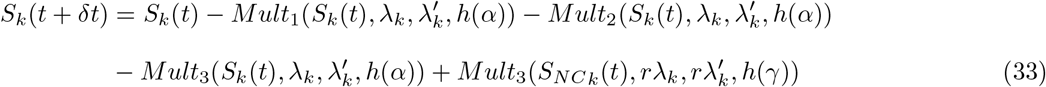

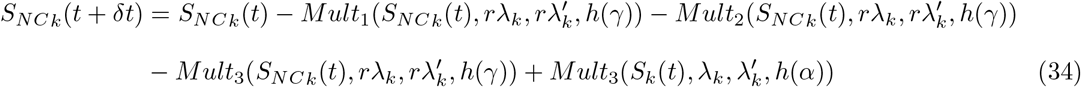

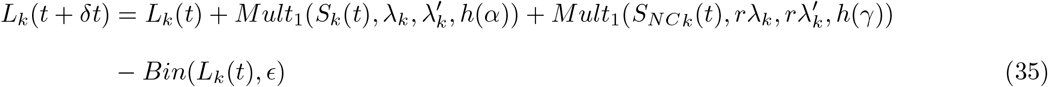

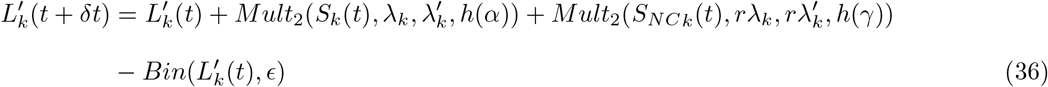

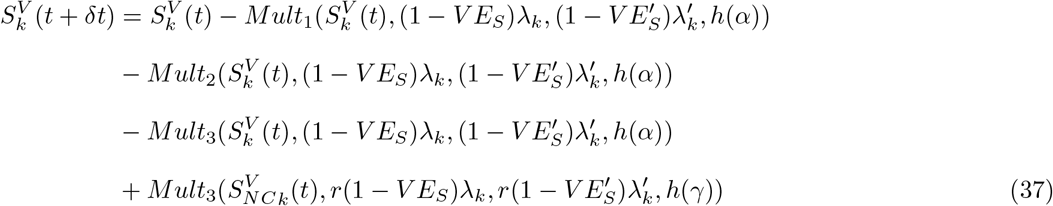

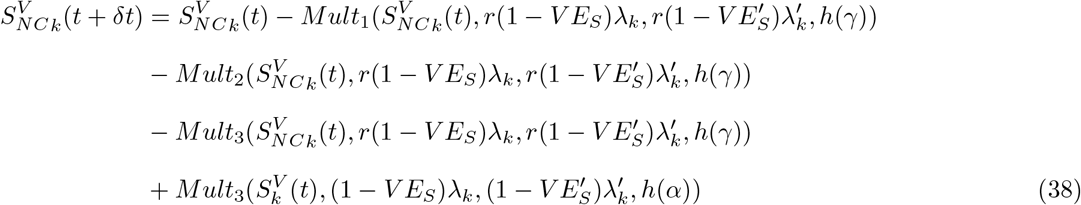

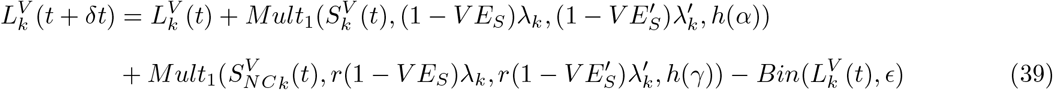

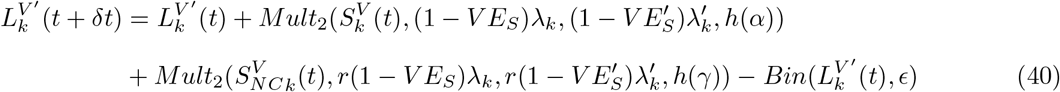

The transitions of the rest compartments are the same as those in the baseline model, described by Eqs 11-30.

Models 3 and 4 can be described by Eqs. 37-40, 9-19, 22-25, and 27-30.

### *R*_0_ calculation

We calculate the basic reproductive number *R*_0_ of proposed model using the next generation matrix approach [72]. By definition, *R*_0_ is the reproductive number at the beginning of the epidemics. Although our models include the emergence of a second variant, vaccines, and relaxation of individual behaviours, all of these become relevant only after the start of the epidemic. As such they have no influence on *R*_0_. Thus, we can disregard the compartments related to the second variant, vaccination, and behavioural relaxation when we calculate *R*_0_. We consider only the infected individuals in the compartments *L*_*k*_ and *I*_*k*_. The deterministic equations regulating the dynamics of these two compartments are:

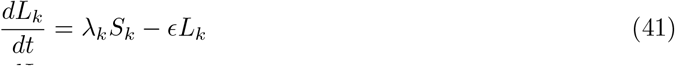

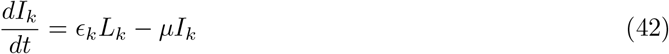

We have *K* = 16 age groups, thus both Eqs. 41 and 42 contains *K* equations for different age groups. We describe these 2*K* equations in matrix form:

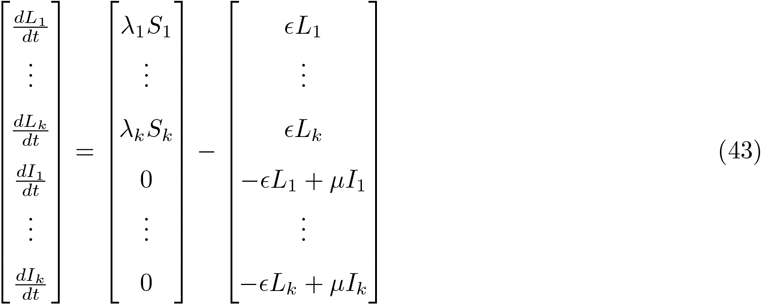

where 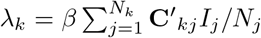 is the force of infection of the original strain, and the contact matrix **C**^′^ accounts for the change in contacts induced by NPIs. We further denote Eq. 43 as

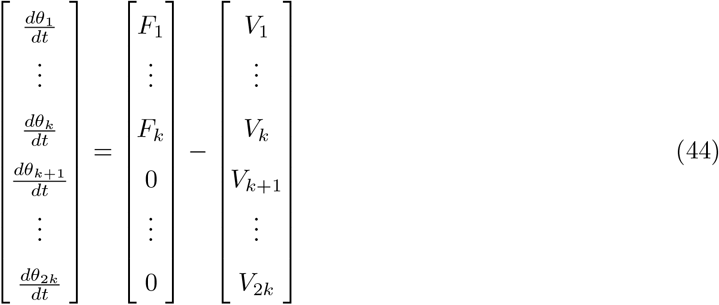

For age group *κ*, we consider the disease free equilibrium (DFE), defined as (*S*_*κ*_, *L*_*κ*_, *I*_*κ*_, *R*_*κ*_)=(*N*_*κ*_, 0, 0, 0). Next, we define two matrices: 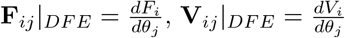. Considering the DFE, we write down **F** and **V** as follows.

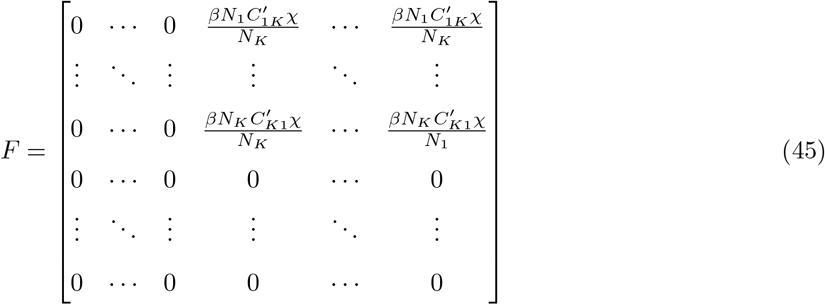

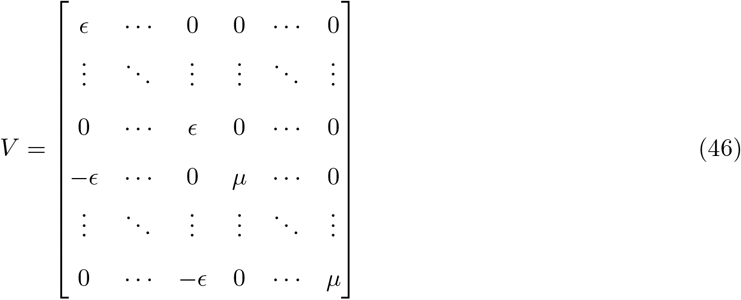

The reproductive number *R*_0_ is defined as *ρ*(**FV**^−**1**^), where *ρ*(·) represents the spectral radius. Then, we write **F** and **V**^−1^ in blocks and we compute **V**^−**1**^ as follows:

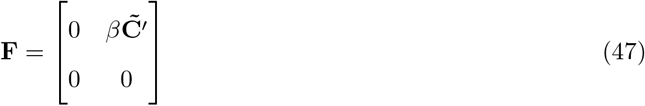

where 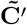 indicates the adjusted contact matrix 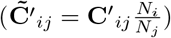 with a size of *K* × *K*, and 0 indicates a *K* × *K* matrix with all zero elements.

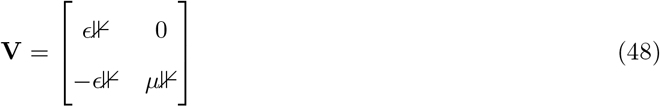

where ⊮ indicates a *K* × *K* identity matrix. Then we compute **V**^−**1**^

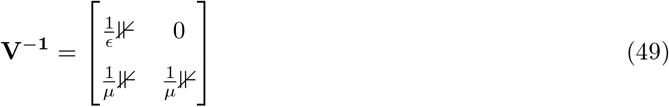

Then, we obtain **FV**^−**1**^

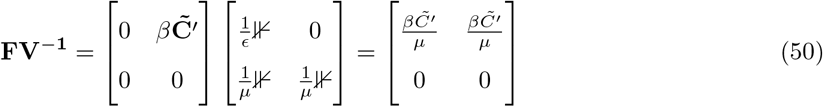

Finally, we are left with finding the spectral radius of **FV**^−**1**^ (i.e., finding its largest eigenvalue). The eigenvalue problem can be written as *det*(**FV**^−**1**^ − *λ*⊮) = 0. Given the structure of **FV**^−**1**^, and since we are interested in non-trivial solutions (λ not equal to 0), the problem reduces to:

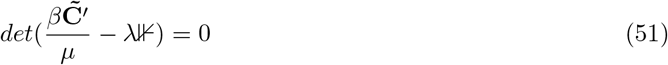

Therefore, we obtain 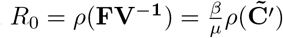.

## 1 Demographics of regions considered

Here, we provide more information about the demographic profiles of the four regions under study.

### Contact matrices

We first present the contact patterns among different age groups. In Fig. 6, we show the pre-Pandemic contact matrices of each region in four settings (i.e., home, school, work places, and other places) sourced from Ref. [65]. Across settings and regions we use the same colour scale for comparison. The plot shows that, across the board, the four regions exhibit similar contact patterns. In detail, the contacts at home show a higher intensity along the diagonal. Contacts at school are the most intense compared to the other three settings, where interactions are mainly among children and adolescents (i.e., age brackets 0 − 4, 5 − 94, 10 − 14, 15 − 19). This is followed by contacts at other places among the young population (i.e., 10 − 14, 15 − 19, 20 − 24, 25 − 29). Contacts at workplaces are less intense than in schools and other places, and are reported mainly among the middle-aged population. By considering contacts across all four settings, we obtain the aggregated contact matrices shown in the last column. The darker colour of the diagonal suggests that within-group interactions (i.e., among people in the same age group) are significantly more frequent than interactions across different age groups. Additionally, teenagers and young adults tend to have more contacts than the elderly population. To compare the contact matrices of the four regions, we compute the spectral radius of the aggregated contact matrix. We find that the spectral radius is 15.2 in British Columbia, 17.0 in Lombardy, 11.7 in London, and 19.5 in São Paulo. As discussed in the previous section, this indicates that, in the context of epidemic spreading, for a given disease London would feature the lowest *R*_0_ while São Paulo the largest.

**Fig 6.**
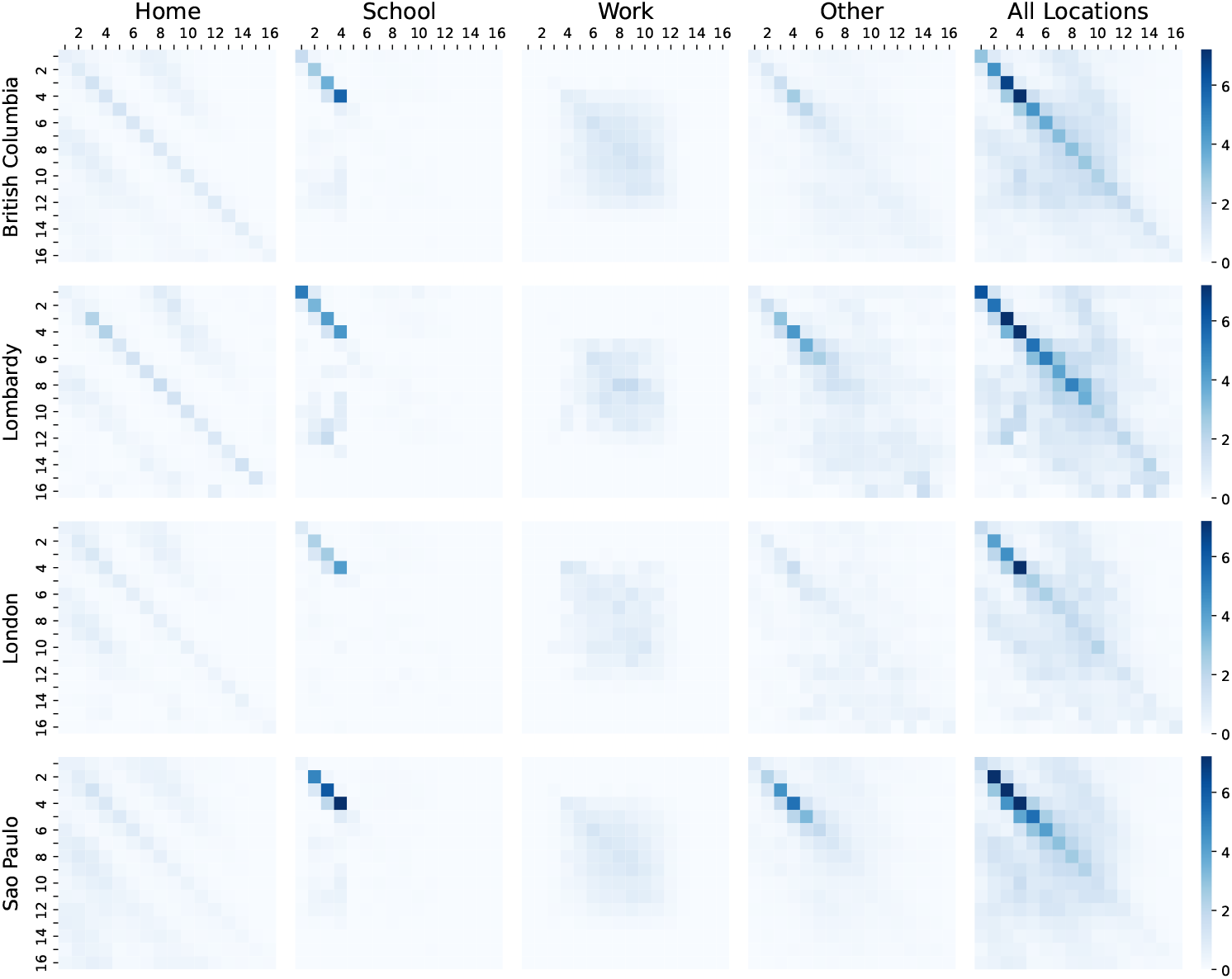
Contact matrices of the four regions. We show the four layers (i.e., home, school, workplace, and other places) of contact matrices between 16 age groups in each region (British Columbia, Lombardy, London, São Paulo). The numbers shown on the top and left of the matrices indicates age groups (e.g., 2 represents age group 5 − 9 and 16 represents age group 75+). The last column shows aggregated contact matrices considering all interaction settings.

### Contact intensity and population distribution

In Fig. 7 we show the pre-Pandemic contact intensity (panel A) and age distribution (panel B) for each age group in the four regions. Contact intensity is computed by summing each column of the overall contact matrix.

**Fig 7.**
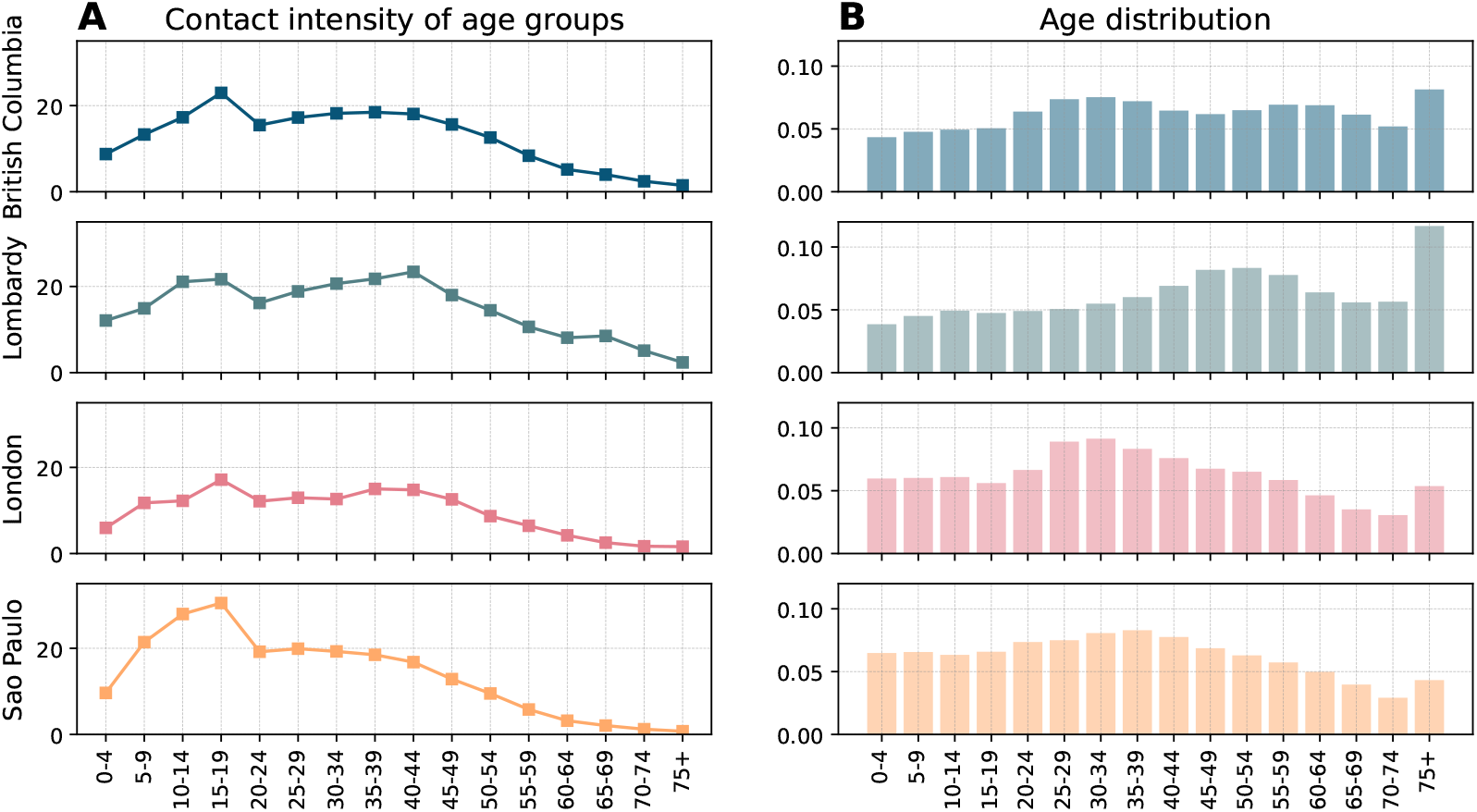
Contact intensity and demographics. A) Contact intensity across age groups in the four regions. Contact intensity is computed by summing each column of the contact matrix. B) Age distributions across the four regions with a 5-year bracket except for the last 75+ group.

Younger age groups, particularly children and adolescents, exhibit the highest contact intensity across all regions, while the elderly population (i.e., 65+) the lowest. In detail, contact intensity peaks in the 15 − 19 age group, followed by a general decline with age with small fluctuations including a slight increase in the 35 − 39 age group (except in São Paulo, where the decline is consistent). A downward trend in contact intensity is observed from the 40 − 44 age group onwards in all regions.

Fig. 7-B shows the population distributions of the four regions in 2021 [73–76]. In British Columbia, the population is relatively evenly distributed across most age groups, with small double peeks in the 30 − 34 to 55 − 59 age groups. Lombardy shows a gradual increase in population from the 30 − 34 age group to the 50 − 54 age group. The population distribution in London peaks in the 30 − 34 age group, suggesting a substantial proportion of young adults. São Paulo shows a peak in the 35 − 39 age group. Comparing the population distributions of the four regions, British Columbia and Lombardy display signs of ageing and also display smaller populations in younger age categories (i.e., 0 − 4, 5 − 9 age groups).

## Impact of vaccines and NPIs

### Timing of vaccination impact

In the context of the no-vaccination counterfactual with the baseline model, we investigate when vaccinations start to have a macroscopic impact. To this end, we compute the time when the weekly deaths with/without vaccinations begin to diverge by at least 1% (i.e., the weekly relative difference of deaths exceeds this threshold). The results are shown in Fig. 8. The grey lines mark the start of vaccinations, and the red lines denote the week in which the vaccination begins to have impact. We find that the vaccination impact started 18 weeks after the first rollout in British Columbia, 6 weeks in Lombardy, 8 weeks in both London and São Paulo. British Columbia experienced a much lower epidemic burden, thus the macroscopic impact of vaccination shows later than the other regions. Notably, the weekly RDD drops to 0 after 2021-15 in London as there are no deaths (the denominator becomes 0).

**Fig 8.**
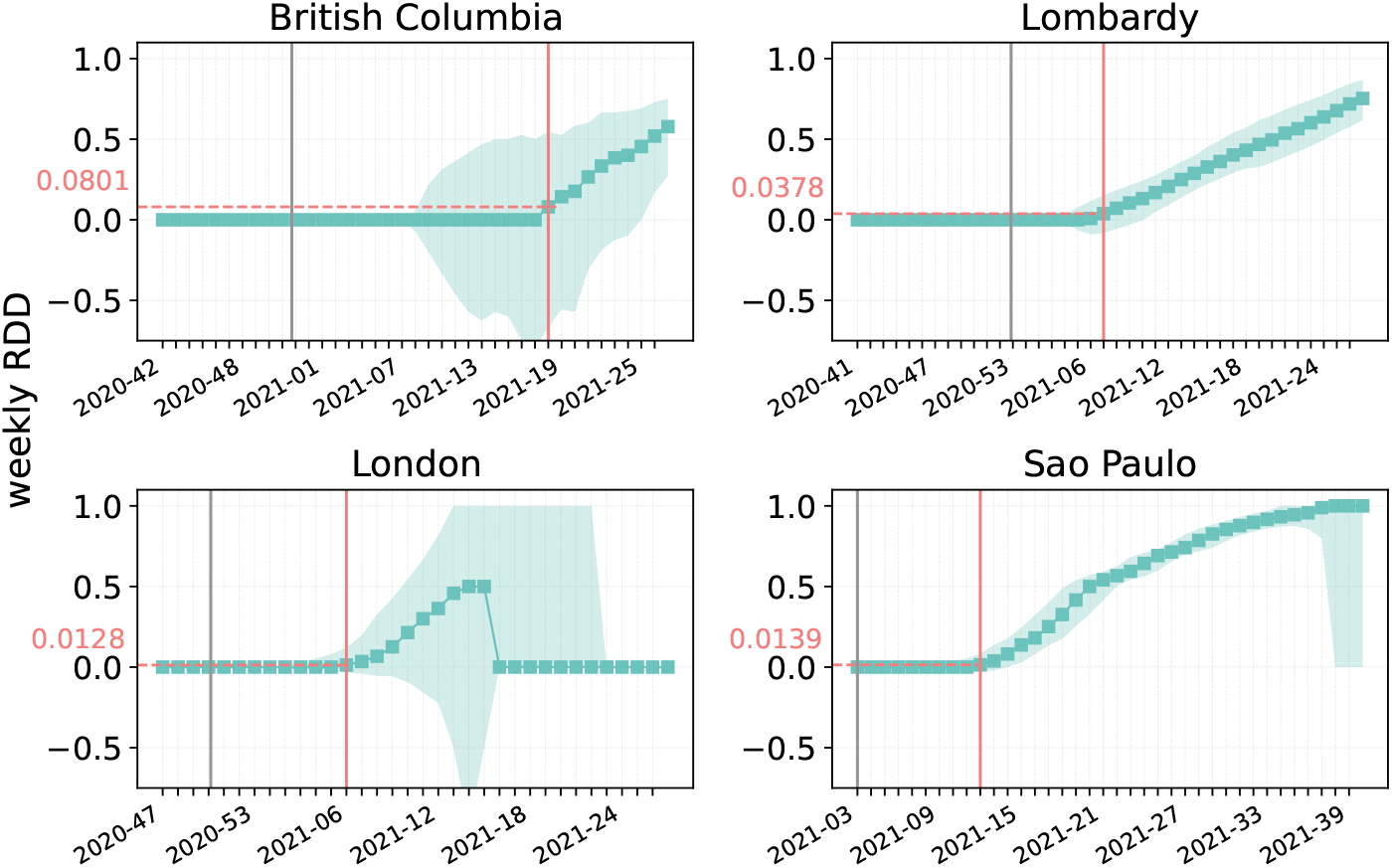
Weekly relative deaths difference in counterfactual scenarios without vaccinations. The fraction of deaths averted by vaccination each week with respect to an equivalent model without vaccinations. The grey lines mark the start of vaccinations while the red lines mark the week in which the relative difference of deaths is larger than 0.01 for the first time (0.0801 in British Columbia, 0.0079 in Lombardy, 0.0128 in London, 0.0139 in São Paulo). We show the results considering 1000 stochastic trajectories, median and 90% confidence interval.

### Influence of NPIs on peak deaths

We analyze the impact of non-pharmaceutical interventions (NPIs) on deaths peak by calculating the fold increase in the peak intensity in a scenario without NPIs compared to the original model with NPIs. In doing so, we maintain vaccination in both scenarios. We first show the death trajectories with/without NPIs in Fig. 9. The peak of deaths without NPIs (red lines) is much higher than with NPIs (blue lines). The peak without NPIs also comes earlier (in terms of the median trajectory) compared to those with NPIs in Lombardy and London (3 weeks earlier) and São Paulo (5 weeks earlier). Next we calculate the fold increase in peak deaths of 1000 stochastic trajectories in Fig. 10. Removing NPIs would have resulted in 5.3 (90% CI: [3.3, 9.4]) times higher in British Columbia, 8.8 [6.8, 11.9] times higher in Lombardy, 6.7 [5.5, 8.2] times higher in London, and 4.7 [3.9, 5.7] times higher in São Paulo times higher peaks. Sao Paulo reports the lowest fold in peak deaths as it reports the least stringent NPIs compared to the other three regions. Lombardy reports the highest fold in peak death, though London adopted the most stringent NPIs. This may be due to a demographic factor. Indeed, Lombardy has a larger proportion of senior population, associated with a higher infection fatality rate due to increased vulnerability to severe outcomes.

**Fig 9.**
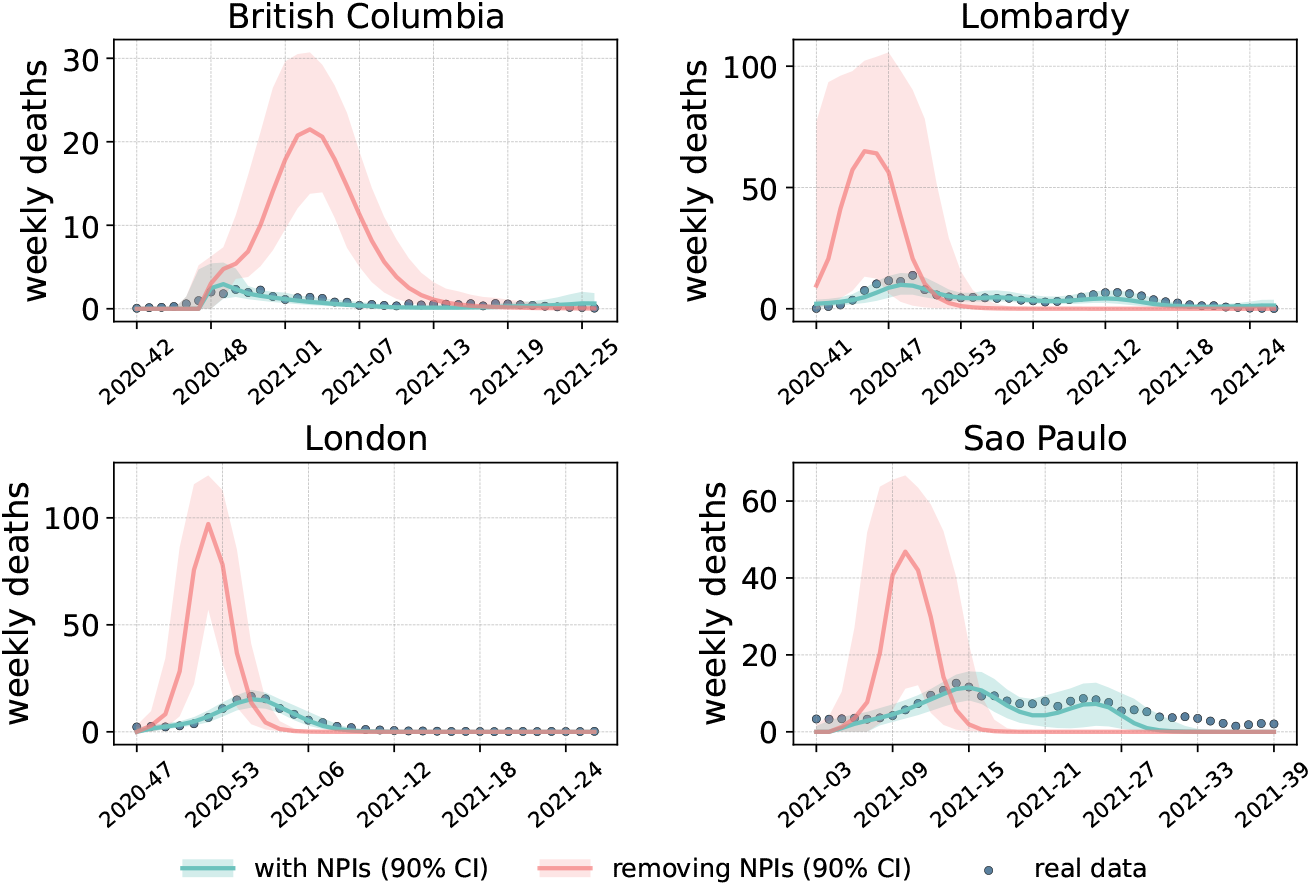
Comparison of weekly death trajectories with/without NPIs. Calibrated weekly death trajectories (weekly deaths per 100, 000) of the baseline model (denoted by blue lines) and in a counterfactual scenario where NPIs are removed (denoted by red lines).

**Fig 10.**
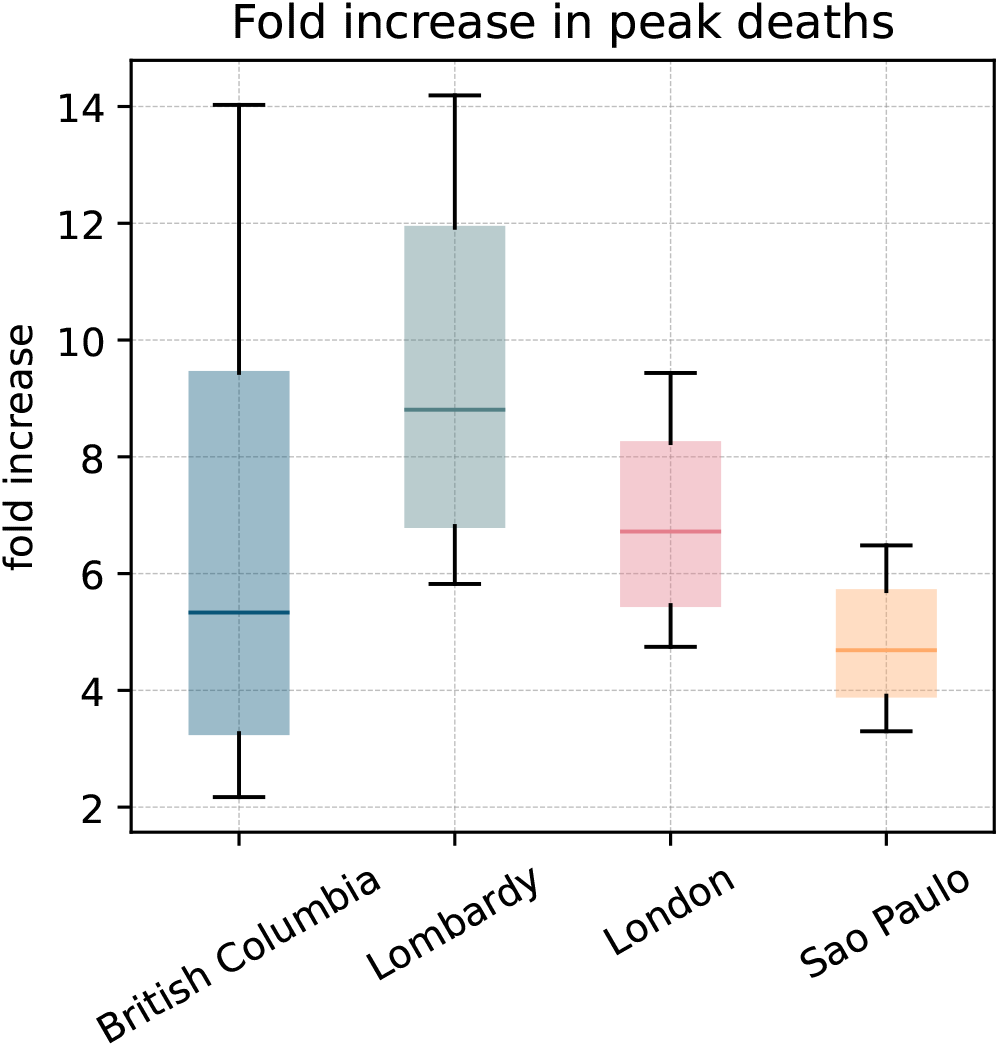
Fold increase in peak deaths intensity in counterfactual scenarios without NPIs. The increase fold of peak deaths in a scenario where NPIs are removed with respect to an equivalent case with NPIs. The box plots show the results of averted deaths considering 1000 stochastic trajectories in each region.

### Relative difference of infection in counterfactual scenarios without vaccination or NPIs

In the main text, we presented the relative deaths difference (i.e., RDD) with/without vaccination, NPIs, and behavioural relaxation. Here, we show the analogous results for the relative difference in the number of infected (i.e., RDI). This quantity is defined as:

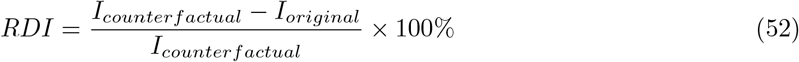

where *I*_*original*_ and *I*_*counterfactual*_ are the total number of simulated infections in the original model and in the counterfactual scenario respectively.

For example, in the case of NPIs *I*_*counterfactual*_ is computed considering estimates from a model without them. Instead, *I*_*original*_ is the corresponding value in matched model with NPIs.

The RDIs in a scenario without vaccination are shown in Fig. 11-A. We compute the median of RDI with a 90% confidence interval considering 1000 stochastic trajectories. The results indicate that vaccination prevented 48.96% (90% CI: [33.69%,58.24%]) of infections in British Columbia, 27.5% ([17.65%,35.68%]) in Lombardy, 0.24% ([0.05%,1.03%]) in London, and 8.59% ([7.14%,13.89%]) in São Paulo. The figures show a similar pattern with the RDDs (shown in the main text) in British Columbia, Lombardy, and London. However, São Paulo shows a different pattern. Indeed, São Paulo has the largest vaccine coverage among the regions and it shows the highest RDD. However, it features a lower RDIs compared to RDDs. São Paulo experienced the Gamma and Delta variants, which reduce the vaccine efficacy against infections significantly (0.65 of Gamma variant and 0.6 of Delta variant).

**Fig 11.**
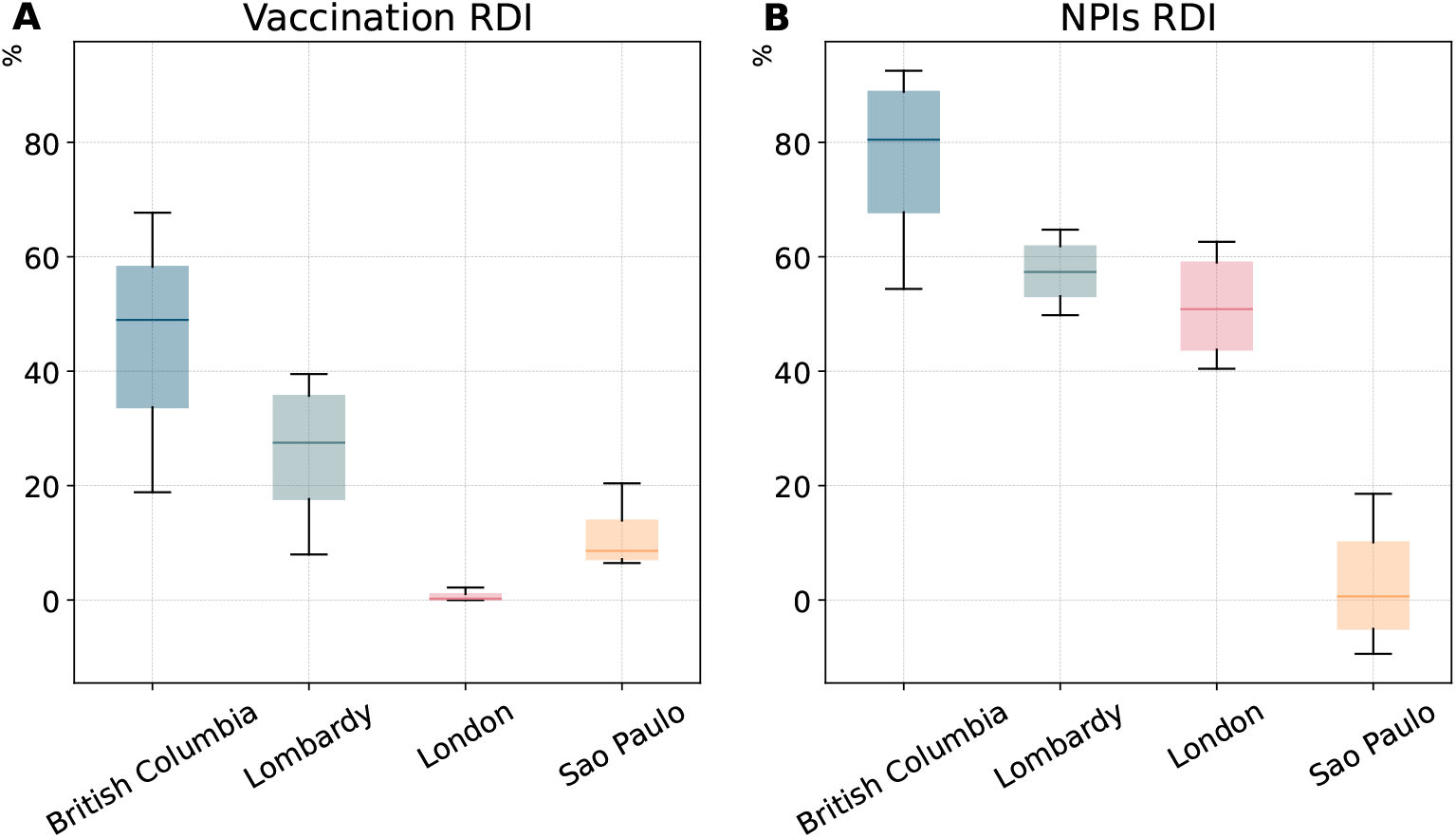
Relative infection difference in counterfactual scenarios without vaccinations and NPIs. Panel A shows the fraction of total infections averted by vaccinations. Panel B shows the fraction of total infections averted by NPIs. The box plots show the results of averted deaths in 1000 stochastic trajectories in each region. The horizontal line within each box marks the median value, while the top and bottom edges correspond to the 0.95 and 0.05 quantiles (90% confidence interval). The whiskers extend to the maximum and minimum values. These estimates are obtained considering the baseline model.

The RDIs of a counterfactual scenario without NPIs are shown in Fig. 11-B. We find 80.46% (90% CI: [67.79%,88.84%]) infections avoided by NPIs in British Columbia, 57.34% ([53.12%,61.83%]) in Lombardy, 50.85% ([43.76%,59.01%]) London in London, and 0.63% ([−5.04%, 10.09%]) in São Paulo. The RDIs of the four regions show similar patterns to the observations for RDDs except for London reporting slightly lower RDIs than Lombardy. This may be explained by a larger population of younger population in London with respect to Lombardy. Besides, by comparing panel A and panel B in Fig. 11, we can conclude that overall, in the first months of vaccines rollout, NPIs averted more infections compared to those prevented by vaccinations. These results highlight one more time the importance of NPIs during the complex initial phases of the vaccination campaign.

### Relative difference of infection in counterfactual scenarios without behavioural relaxation

We also calculate the relative difference of infections (RDIs) considering a counterfactual scenario where we remove the relaxation from the four behavioural models. The results are shown in Fig. 12 and Table 5, displaying the median of RDIs along with 50% CIs. The values of RDI are below 0 in most cases, which means removing behavioural mechanisms leads to fewer infections (see Table 5). The results for RDIs are consistent with the analogous for RDDs shown in the main text.

**Fig 12.**
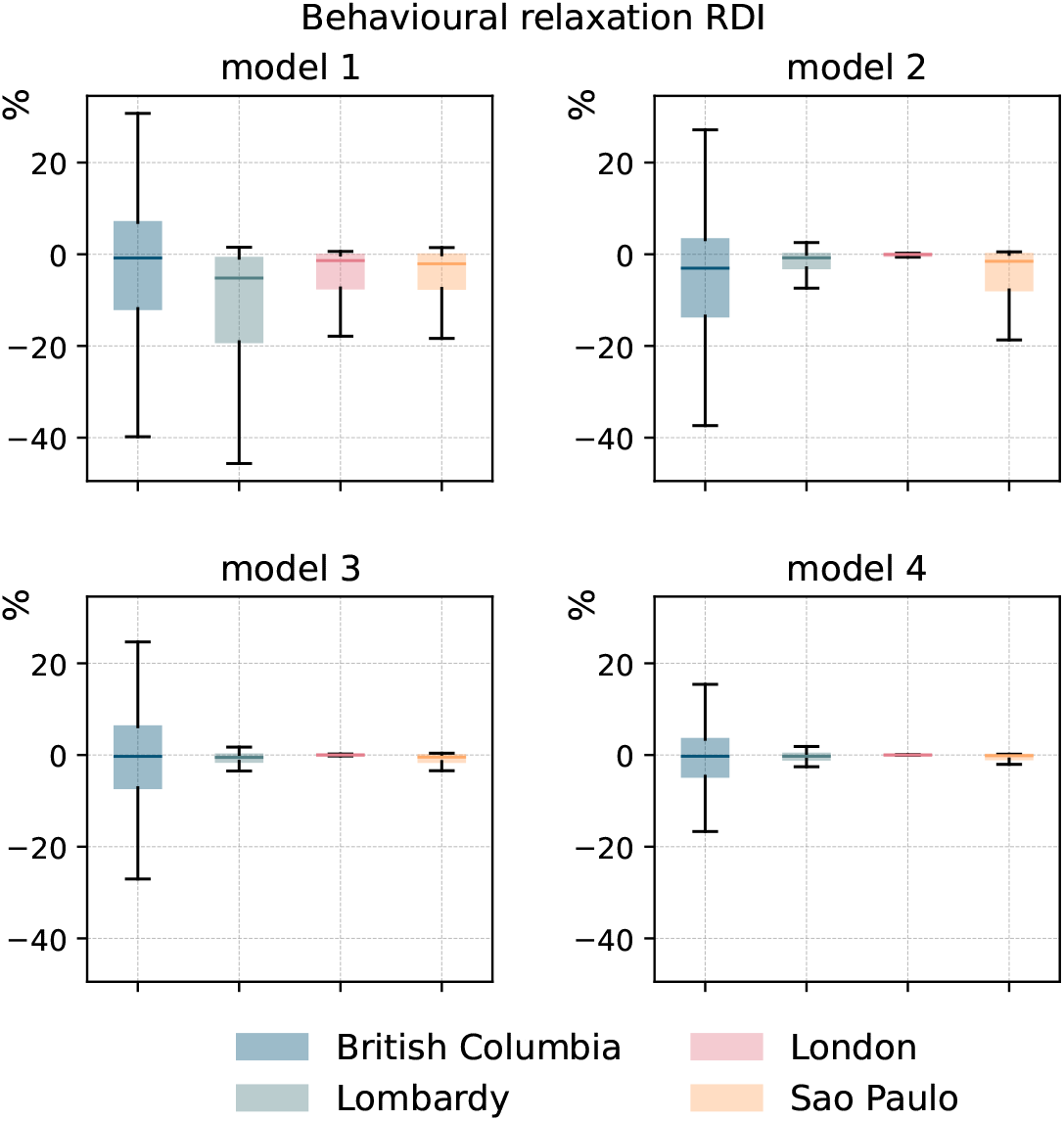
Relative infection difference in counterfactual scenarios without behavioural relaxation. The fraction of total infections averted by behavioural relaxation. The box plots show the results of averted deaths in 1000 stochastic trajectories in each region. The horizontal line within each box marks the median value, while the top and bottom edges correspond to the 0.25 and 0.75 quartiles. The whiskers extend to the maximum and minimum values after removing the outliers that beyond the interquartile range.

**Fig 13.**
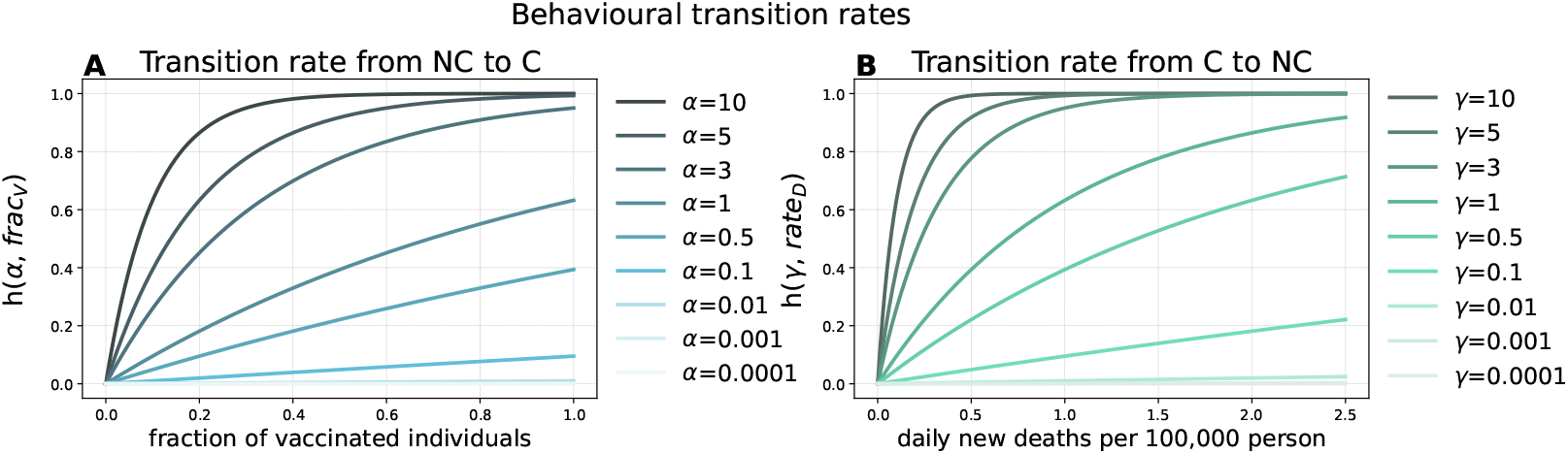
Behavioural transition rates for models 2 and 4. In panel A we plot the behavioural transition rate from non-compliant to compliant denoted by *h*(*α, frac*_*V*_) as a function of the fraction of vaccinated individuals for different values of *α*. In panel B we plot the behavioural transition rate from compliant to non-compliant denoted by *h*(*γ, rate*_*D*_(*t*)) as a function of the fraction of daily new deaths per 100, 000 for different values of *γ*.

### Absolute death differences across counterfactual scenarios

In the main text, we showed the relative deaths difference considering counterfactual scenarios without vaccinations, NPIs, or behavioural relaxation. Here, we show absolute values. We show the medians with 90% confidence interval in Tables 6-8.

**Table 6.**
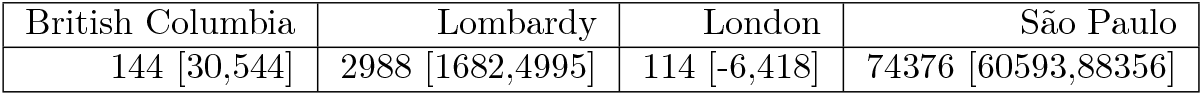
Deaths difference in a counterfactual without vaccinations.

| British Columbia | Lombardy | London | São Paulo |
| --- | --- | --- | --- |
| 144 [30,544] | 2988 [1682,4995] | 114 [-6,418] | 74376 [60593,88356] |

As is shown in Table 6, the absolute deaths difference in counterfactual scenarios without vaccination is small in British Columbia and London with a median of 144 and 114 more deaths without vaccinations than that with vaccination. In contrast, São Paulo shows a large difference with a median of more than 70*K*.

The absolute deaths difference in counterfactual scenarios without NPIs is shown in Table 7. The values are much higher than those of vaccination except a decrease in São Paulo. As mentioned in the main text, in São Paulo we observed the least stringent adoption of NPIs. These results underscore the importance of vaccinations, especially in settings with low adoption of NPIs.

**Table 7.** Deaths difference in a counterfactual without NPIs

| British Columbia | Lombardy | London | São Paulo |
| --- | --- | --- | --- |
| 9266 [7515,11633] | 31337 [26775,37965] | 22256 [18296,27550] | 39448 [30069,54683] |

In Table 8, we show the deaths difference in counterfactual scenarios without behavioural relaxation. Models 1 and 2 in São Paulo, lead to larger number of deaths. In general, the impact of behavioural relaxation on deaths is much smaller than the impact of vaccination and NPIs in each region.

**Table 8.** Death difference in a counterfactual without behavioural relaxation.

|  | model 1 | model 2 | model 3 | model 4 |
| --- | --- | --- | --- | --- |
| British Columbia | -5 [-53,42] | -1 [-37,31] | 4 [-34,39] | 1 [-23,25] |
| Lombardy | -664 [-2384,-120] | -94 [-261,33] | -56 [-166,51] | -22 [-124,61] |
| London | -178 [-746,-35] | -19 [-77,28] | -1 [-48,49] | 0 [-21,22] |
| Sao Paulo | -17399 [-31562,-5540] | -3508 [-15659,-158] | -702 [-1897,-144] | -318 [-1087,20] |

## Impact of behavioural relaxation

### Behavioural transition rate

In model 1 and 3 the transitions towards non-compliance and those back to compliance happen at constant rates, *α* and *γ* respectively. Instead, in models 2 and 4 these transitions are proportional to the fraction of vaccinated (multiplied by *α*) and deaths per 100, 000 (multiplied by *γ*). To better understand these varying transition rates, in Fig. 1-A we plot, for different values of *α*, the transition rates from compliant to non-compliant compartments as a function of the fraction of vaccinated individuals. We denote this rate as 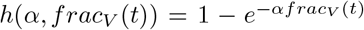, where *frac*_*V*_ (*t*) is the fraction of vaccinated individuals of the total population. Similarly, in Fig. 1-B we consider a range of *γ* values and plot the transition rates from compliant back to non-compliant as a function of daily new deaths per 100, 000 denoted by 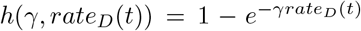, where *rate*_*D*_(*t*) represents daily death rate (deaths per 100, 000).

The results in Fig. 1-A show that a higher fraction of vaccinated individuals leads to an increased transition rates towards non-compliance. As *α* increases, the transition rate *h*(*α, frac*_*V*_) rises more sharply. This means that, for a given fraction of vaccinated individuals, higher values of *α* result in a greater shift towards non-compliance. For example, when *α* = 0.1, the transition rate is only 0.1 even when the fraction of vaccinated individuals reaches its maximum (1). In contrast, when *α* = 10, the transition rate reaches 1 when only 40% of the population is vaccinated. A similar trend is observed in Fig. 1-B where higher daily death rates lead to higher transition rates from non-compliance back to compliance. Furthermore, higher values of *γ* result in stronger responses to the epidemic’s severity. For example, when *γ* = 0.1, the transition rate is 0.2 even when the daily death rate is quite high (2). In contrast, when *γ* = 10, the transition rate reaches the maximum when the death rate is smaller (0.5).

### Fraction of non-compliant individuals as function of time

To have a more intuitive understanding on the four behavioural mechanisms, in Fig. 14 we plot the fraction of non-compliant individuals as function of time for the four models. To this end, we consider 1000 simulations obtaining sampling the posterior distribution of each model. We then compute the fraction of non-compliant individuals for each. We show the median trajectory with 90% confidence intervals. Overall, we observe that model 1 result in the largest fraction of non-compliant individuals in the Lombardy and Sao Paulo. We also note that the models 1 and 2 show the largest variability in terms of their confidence intervals. Besides, in the case of Sao Paulo, the median of trajectory of the fraction of NC of model 1 quickly raises to about 0.4 of the population, being stable for a while then showing a slow decrease. As shown in the main text, this is consistent with the large RDD by behavioural relaxation for this model and location.

**Fig 14.**
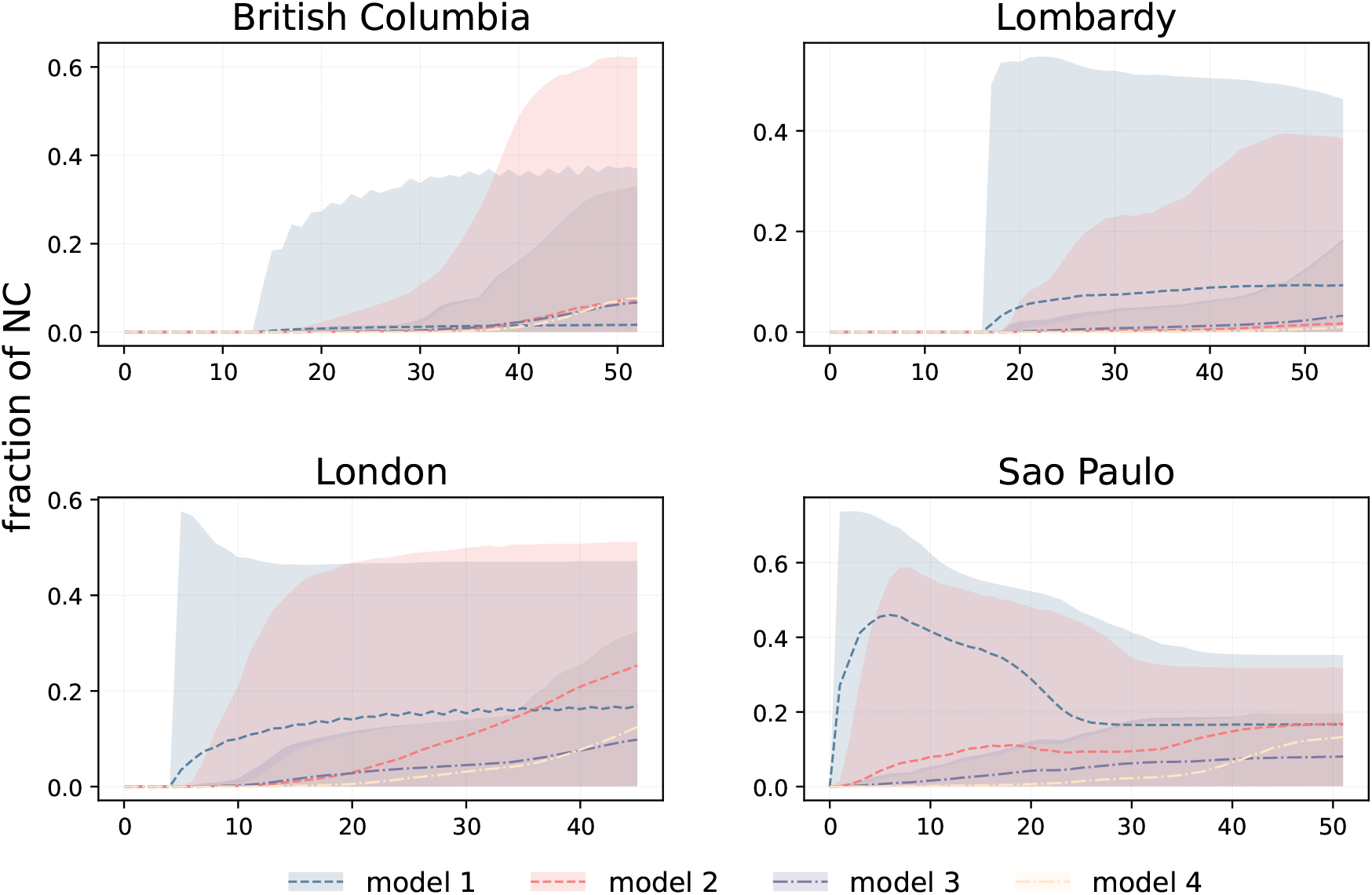
Fraction of non-compliant individuals as function of time. We plot the fraction of non-compliant individuals of the four behavioural models (models 1–4) in each region. The fractions are computed considering 1000 sampled trajectories. The dotted lines represent the median fractions, with shaded areas indicating the 90% confidence intervals.

## Model calibration

### AIC/BIC scores of models

In Table 9 we report the AIC scores of the five models. Smallest scores indicates a better performance of a model. As is shown, the baseline reports the smallest scores in the three regions (British Columbia, Lombardy, and London) out of four, while only one behavioural model (model 2) reports the smallest AIC score in São Paulo.

**Table 9.**
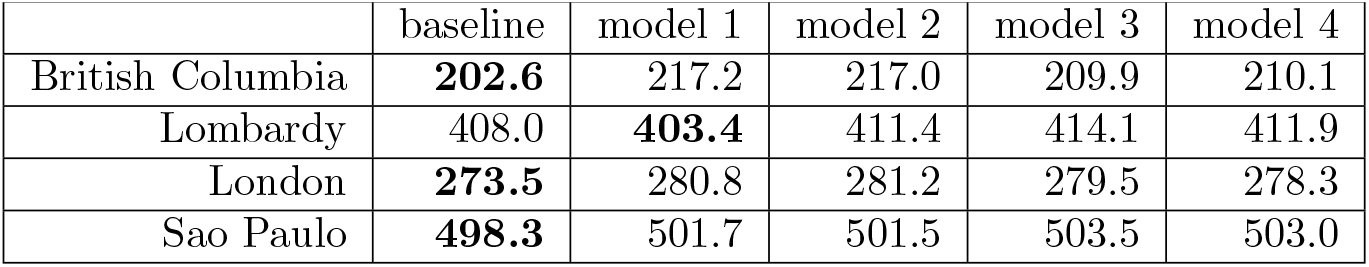
AIC scores of models.

|  | baseline | model 1 | model 2 | model 3 | model 4 |
| --- | --- | --- | --- | --- | --- |
| British Columbia | **202.6** | 217.2 | 217.0 | 209.9 | 210.1 |
| Lombardy | 408.0 | **403.4** | 411.4 | 414.1 | 411.9 |
| London | **273.5** | 280.8 | 281.2 | 279.5 | 278.3 |
| Sao Paulo | **498.3** | 501.7 | 501.5 | 503.5 | 503.0 |

Similarly, we show the BIC scores of the five models in Table 10. Smallest scores indicates a better performance of a model. As is shown, the baseline reports the smallest scores in all the regions, though in Lombardy model 1 has very similar BIC scores to the baseline model.

**Table 10.** BIC scores of models.

|  | baseline | model 1 | model 2 | model 3 | model 4 |
| --- | --- | --- | --- | --- | --- |
| British Columbia | **211.4** | 230.4 | 230.2 | 223.1 | 223.3 |
| Lombardy | **418.0** | 418.4 | 426.4 | 429.0 | 426.9 |
| London | **280.8** | 292.5 | 292.9 | 291.2 | 290.0 |
| Sao Paulo | **509.2** | 517.3 | 517.1 | 519.1 | 518.5 |

**Table 11.**
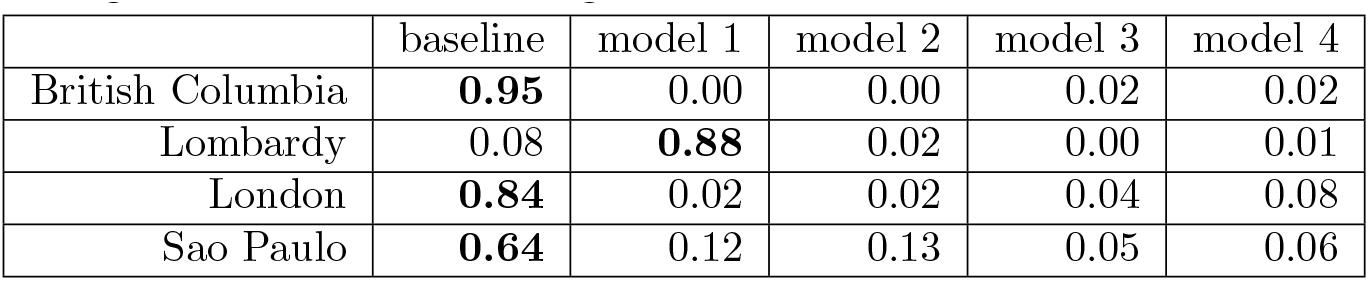
AIC weights of models of removing the last 1 week.

|  | baseline | model 1 | model 2 | model 3 | model 4 |
| --- | --- | --- | --- | --- | --- |
| British Columbia | **0.95** | 0.00 | 0.00 | 0.02 | 0.02 |
| Lombardy | 0.08 | **0.88** | 0.02 | 0.00 | 0.01 |
| London | **0.84** | 0.02 | 0.02 | 0.04 | 0.08 |
| Sao Paulo | **0.64** | 0.12 | 0.13 | 0.05 | 0.06 |

**Table 12.** AIC weights of models of removing the last 2 weeks.

|  | baseline | model 1 | model 2 | model 3 | model 4 |
| --- | --- | --- | --- | --- | --- |
| British Columbia | **0.95** | 0.00 | 0.00 | 0.02 | 0.02 |
| Lombardy | 0.08 | **0.89** | 0.02 | 0.00 | 0.01 |
| London | **0.84** | 0.02 | 0.02 | 0.04 | 0.08 |
| Sao Paulo | **0.64** | 0.12 | 0.13 | 0.05 | 0.06 |

**Table 13.**
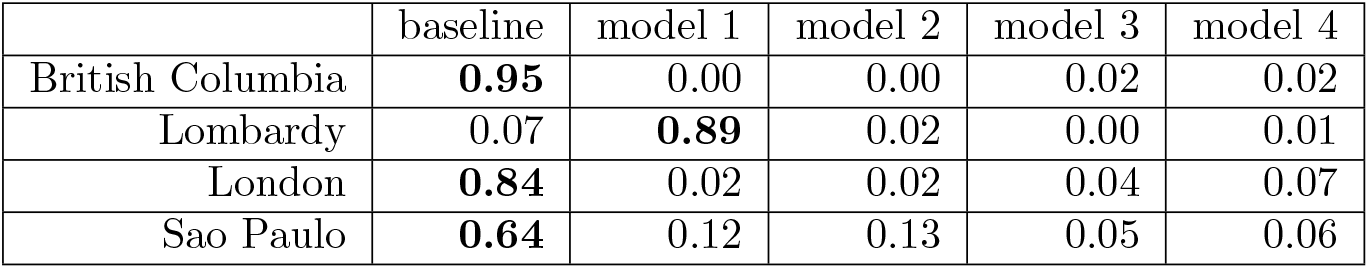
AIC weights of models of removing the last 3 weeks.

|  | baseline | model 1 | model 2 | model 3 | model 4 |
| --- | --- | --- | --- | --- | --- |
| British Columbia | **0.95** | 0.00 | 0.00 | 0.02 | 0.02 |
| Lombardy | 0.07 | **0.89** | 0.02 | 0.00 | 0.01 |
| London | **0.84** | 0.02 | 0.02 | 0.04 | 0.07 |
| Sao Paulo | **0.64** | 0.12 | 0.13 | 0.05 | 0.06 |

**Table 14.** AIC weights of models of removing the last 4 weeks.

|  | baseline | model 1 | model 2 | model 3 | model 4 |
| --- | --- | --- | --- | --- | --- |
| British Columbia | **0.95** | 0.00 | 0.00 | 0.02 | 0.02 |
| Lombardy | 0.08 | **0.88** | 0.02 | 0.00 | 0.01 |
| London | **0.84** | 0.02 | 0.02 | 0.04 | 0.07 |
| Sao Paulo | **0.65** | 0.11 | 0.13 | 0.05 | 0.06 |

### Sensitivity analysis of AIC weight

We re-compute the AIC weights removing the last 1, 2, 3, 4 week(s) in the trajectories. The results are shown in Tables. 11-14. The models’ performance in terms of AIC weights is robust. Model 1 is the most likely model in Lombardy and the baseline in the other three regions in all cases.

### Sensitivity analysis of BIC

We computed also the BIC to measure models’ performance. Similar to AIC, BIC also consider both fit and complexity of models, however, BIC prefers simpler models than AIC as it penalize the complexity more than AIC. The BIC weights of models are shown in Tables 15-19. Table 15 displays the BIC weights computed using complete trajectories, while Tables 16, 17, 18, and 19 show the results of sensitivity tests where we removed the last 1, 2, 3, 4 week(s) from the trajectories respectively. Model 1 in Lombardy does not emerge as the most likely when considering BIC weights for either the complete trajectories or when excluding the last 1 data point. The difference between model 1 and the baseline is however rather small. When we remove the last 2, 3, 4 points, model 1 in Lombardy becomes the most likely model. In general, the results are robust across AIC and BIC weights and different temporal horizons of data.

**Table 15.**
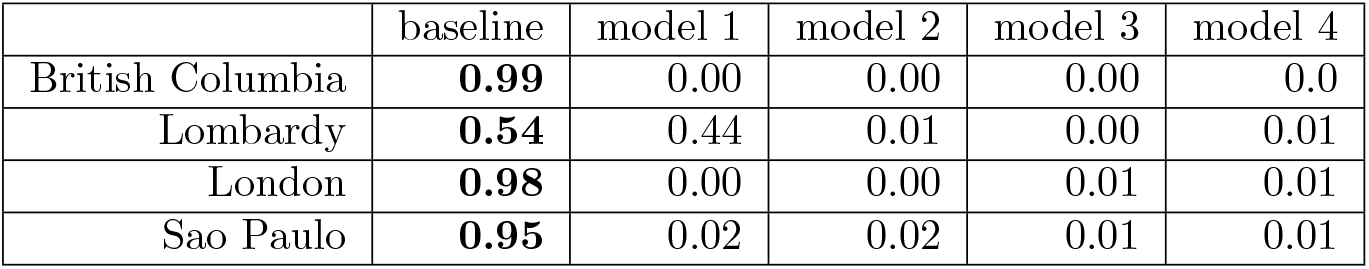
BIC weights of models.

**Table 16.** BIC weights of models of removing the last 1 week.

|  | baseline | model 1 | model 2 | model 3 | model 4 |
| --- | --- | --- | --- | --- | --- |
| British Columbia | **0.99** | 0.00 | 0.00 | 0.00 | 0.00 |
| Lombardy | **0.51** | 0.47 | 0.01 | 0.00 | 0.01 |
| London | **0.98** | 0.00 | 0.00 | 0.01 | 0.01 |
| Sao Paulo | **0.95** | 0.02 | 0.02 | 0.01 | 0.01 |

**Table 17.** BIC weights of models of removing the last 2 weeks

|  | baseline | model 1 | model 2 | model 3 | model 4 |
| --- | --- | --- | --- | --- | --- |
| British Columbia | **0.99** | 0.00 | 0.00 | 0.00 | 0.00 |
| Lombardy | 0.48 | **0.50** | 0.01 | 0.00 | 0.01 |
| London | **0.98** | 0.00 | 0.00 | 0.01 | 0.01 |
| Sao Paulo | **0.94** | 0.02 | 0.02 | 0.01 | 0.01 |

**Table 18.** BIC weights of models of removing the last 3 weeks

|  | baseline | model 1 | model 2 | model 3 | model 4 |
| --- | --- | --- | --- | --- | --- |
| British Columbia | **0.99** | 0.00 | 0.00 | 0.00 | 0.00 |
| Lombardy | 0.46 | **0.51** | 0.01 | 0.00 | 0.01 |
| London | **0.98** | 0.00 | 0.00 | 0.01 | 0.01 |
| Sao Paulo | **0.94** | 0.02 | 0.02 | 0.01 | 0.01 |

**Table 19.** BIC weights of models of removing the last 4 weeks

|  | baseline | model 1 | model 2 | model 3 | model 4 |
| --- | --- | --- | --- | --- | --- |
| British Columbia | **0.99** | 0.00 | 0.00 | 0.00 | 0.0 |
| Lombardy | 0.48 | **0.50** | 0.01 | 0.00 | 0.01 |
| London | **0.98** | 0.00 | 0.00 | 0.01 | 0.01 |
| Sao Paulo | **0.94** | 0.02 | 0.02 | 0.01 | 0.01 |

The results with removing the last 1 week:

The results with removing the last 2 weeks:

The results with removing the last 3 weeks:

The results with removing the last 4 weeks:

### Posterior distributions of parameters

In this section, we present the posterior distributions of the free parameters in our models computed via the Approximate Bayesian Computation-Sequential Monte Carlo (ABC-SMC). For each region, we plot the median values of the sampled parameters, along with the interquartile range (IQR) spanning from the first to the third quartile. Notably, the number of free parameters in the baseline model differs among regions due to the emergence of the second variant. Specifically, for the baseline model, in British Columbia and Lombardy, there are six free parameters to be calibrated: the reproductive number *R*_0_, the delay time in reporting deaths Δ, the initial fraction of infected individuals *i*_*ini*_, the initial fraction of recovered individuals *r*_*ini*_, the adjustment of the start date of the simulation of epidemic Δ*t*, and the adjustment of the introduction date of a VOC Δ*t*_*var*_. For London we have five free parameters as above, excluding Δ*t*_*var*_, as London experienced only one strain during the simulation period. In São Paulo, seven parameters are calibrated, the above six, as for British Columbia and Lombardy, and additionally the relative transmissibility (*σ*) of the Delta variant compared to the Gamma variant. As mentioned in the main text, this relative transmissibility for Alpha is fixed at 1.5 for British Columbia and Lombardy. Across all regions, the behavioural models (Models 1-4) consistently incorporate three additional behavioural parameters: the behavioural transition parameters *α* and *γ*, as well as the relative infection probability *r* of non-compliant individuals.

The posterior distributions of the four regions are shown in Figs. 15-18. We compare the posteriors across the regions. For the reproductive number *R*_0_, the calibrated values span from 1.0 to 2.5. London exhibits the highest *R*_0_ across all models, due to the circulation is Alpha variant at the beginning time of our simulations. British Columbia and Lombardy instead reports lower ranges of *R*_0_, corresponding to the circulation of the wild type in these two regions. The delay in reporting deaths (Δ) ranges from 23 to 64 days across the four regions. British Columbia shows a longer delay. In contrast, Lombardy, shows a shorter delay. This difference may be due to difference in healthcare reporting systems. Regarding the posteriors of initial fraction of infected individuals *i*_*ini*_, Sao Paulo shows the largest fraction with a median of 0.0048 (baseline model) across the four regions. The posteriors of the fraction of recovered *r*_*ini*_ shows the highest value with a median of 0.294 (baseline model) in Lombardy. This is consistent with the fact that Lombardy experienced the highest mortality rate across the four regions till the start date in our simulation. The medians of the adjustment for the start date of simulations (Δ*t*) are 2, 6, 3, 4 weeks in British Columbia, Lombardy, London, and Sao Paulo. These figures suggest the best fit epidemic starting dates are 2, 6, 3, 4 weeks prior to the baseline date *t*∗ set in section Materials and Methods in the main text. The adjustment of the introduction date of a VOC Δ_*var*_ shows a median of 11 in days in British Columbia and Lombardy, while it shows a median of 38 days in São Paulo. Analogously, these figures suggest the best fit dates at which we introduce a second variant with a fraction of 0.01 of infections in British Columbia, Lombardy, and Sao Paulo are 11, 11, 38 days prior to the baseline date 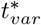 set in section Materials and Methods in the main text.

**Fig 15.**
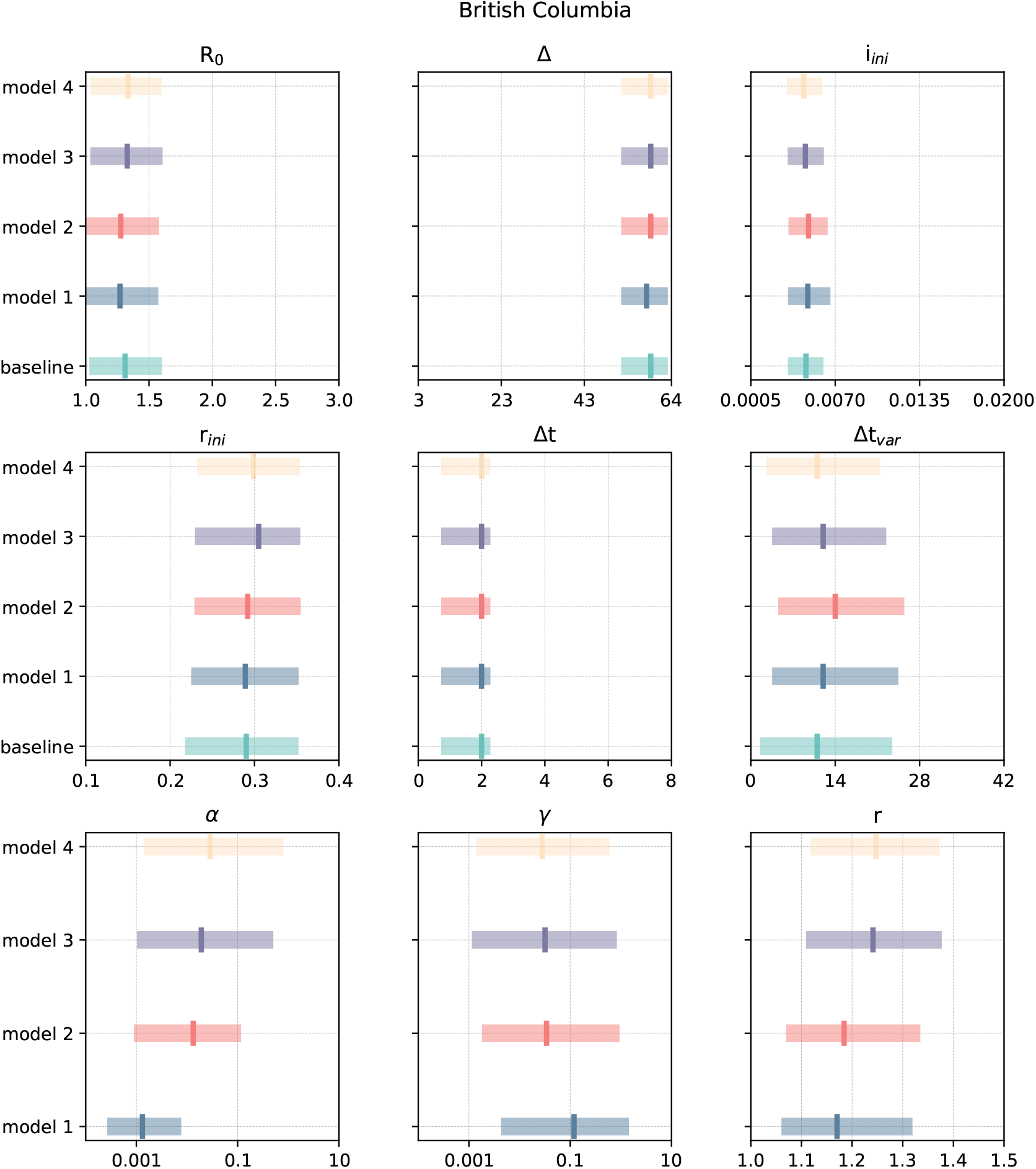
Posterior distributions of calibrated parameters for British Columbia. We plot the median with the range between first quartile and third quartile. The first sixth parameters, *R*_0_, Δ, *i*_*ini*_, *r*_*ini*_, Δ*t*, Δ*t*_*var*_ are calibrated in all the five models. *α, β*, and *r* are behavioural parameters only in behavioural models (models 1-4).

**Fig 16.**
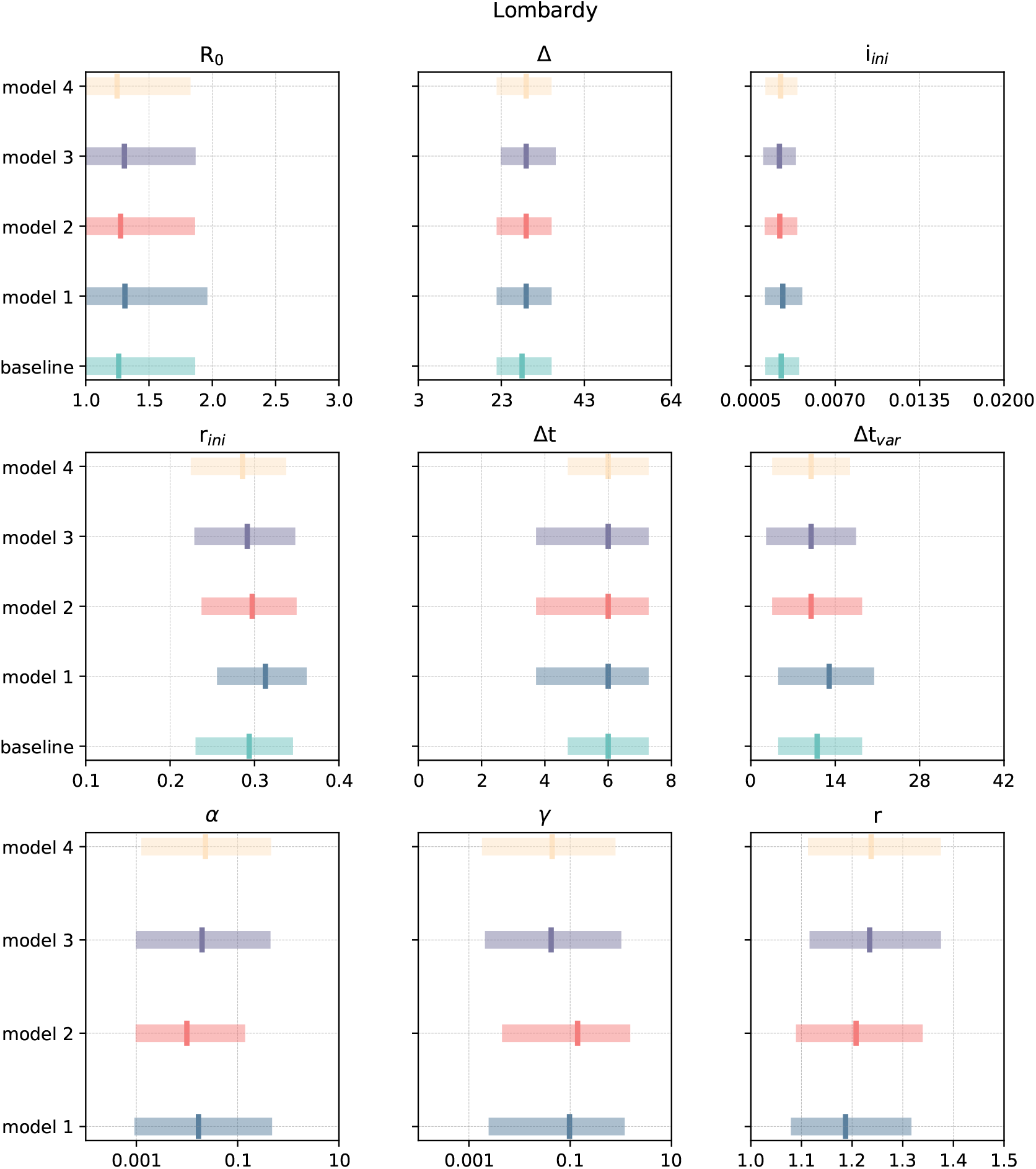
Posterior distributions of calibrated parameters for Lombardy. We plot the median with the range between the first quartile and the third quartile. The first sixth parameters, *R*_0_, Δ, *i*_*ini*_, *r*_*ini*_, Δ*t*, Δ*t*_*var*_ are calibrated in all the five models. *α, β*, and *r* are behavioural parameters only in behavioural models (models 1-4).

**Fig 17.**
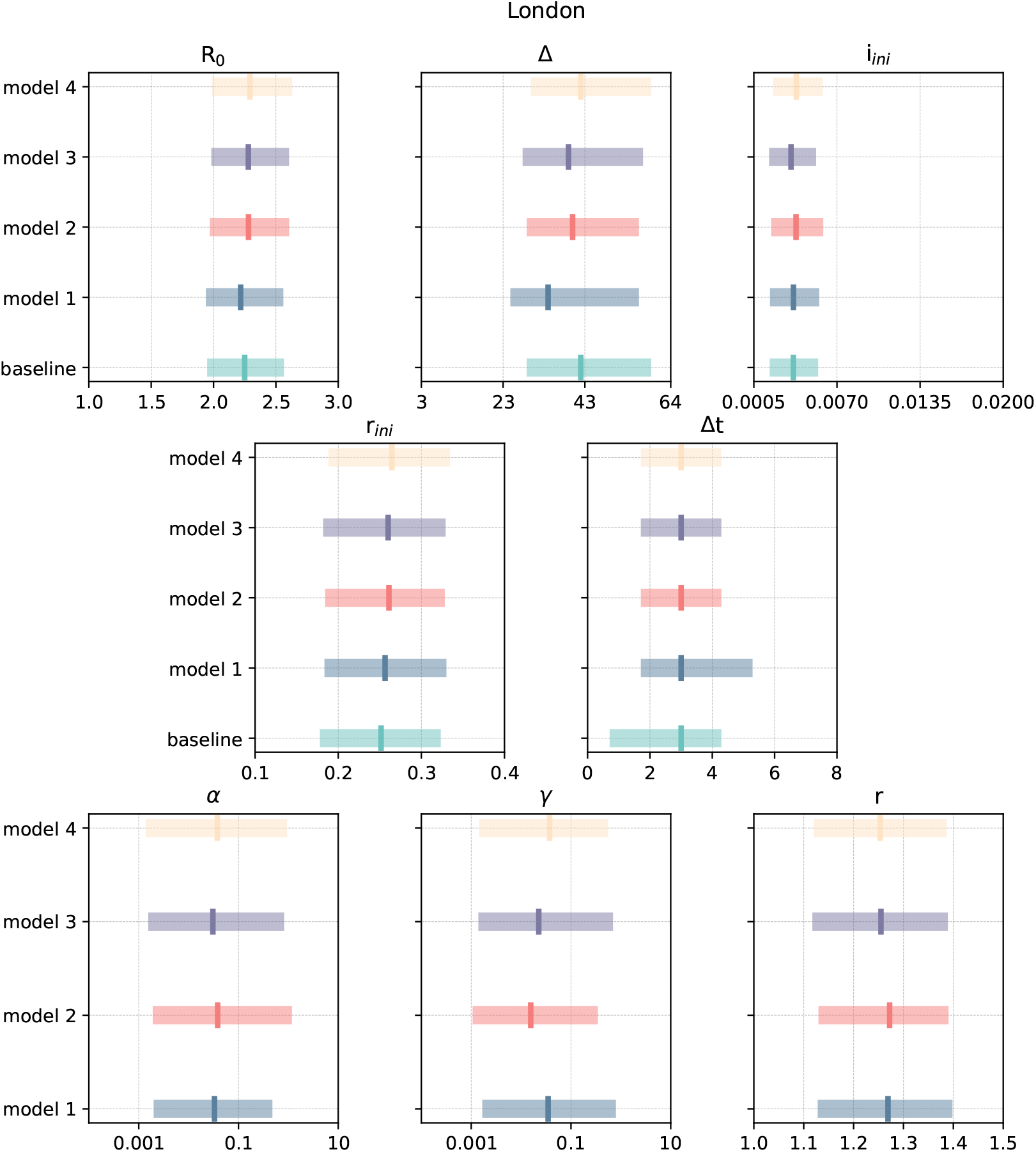
Posterior distributions of calibrated parameters for London. We plot the median with the range between the first quartile and the third quartile. The first sixth parameters, *R*_0_, Δ, *i*_*ini*_, *r*_*ini*_, Δ*t* are calibrated in all the five models. *α, β*, and *r* are behavioural parameters only in behavioural models (models 1-4).

**Fig 18.**
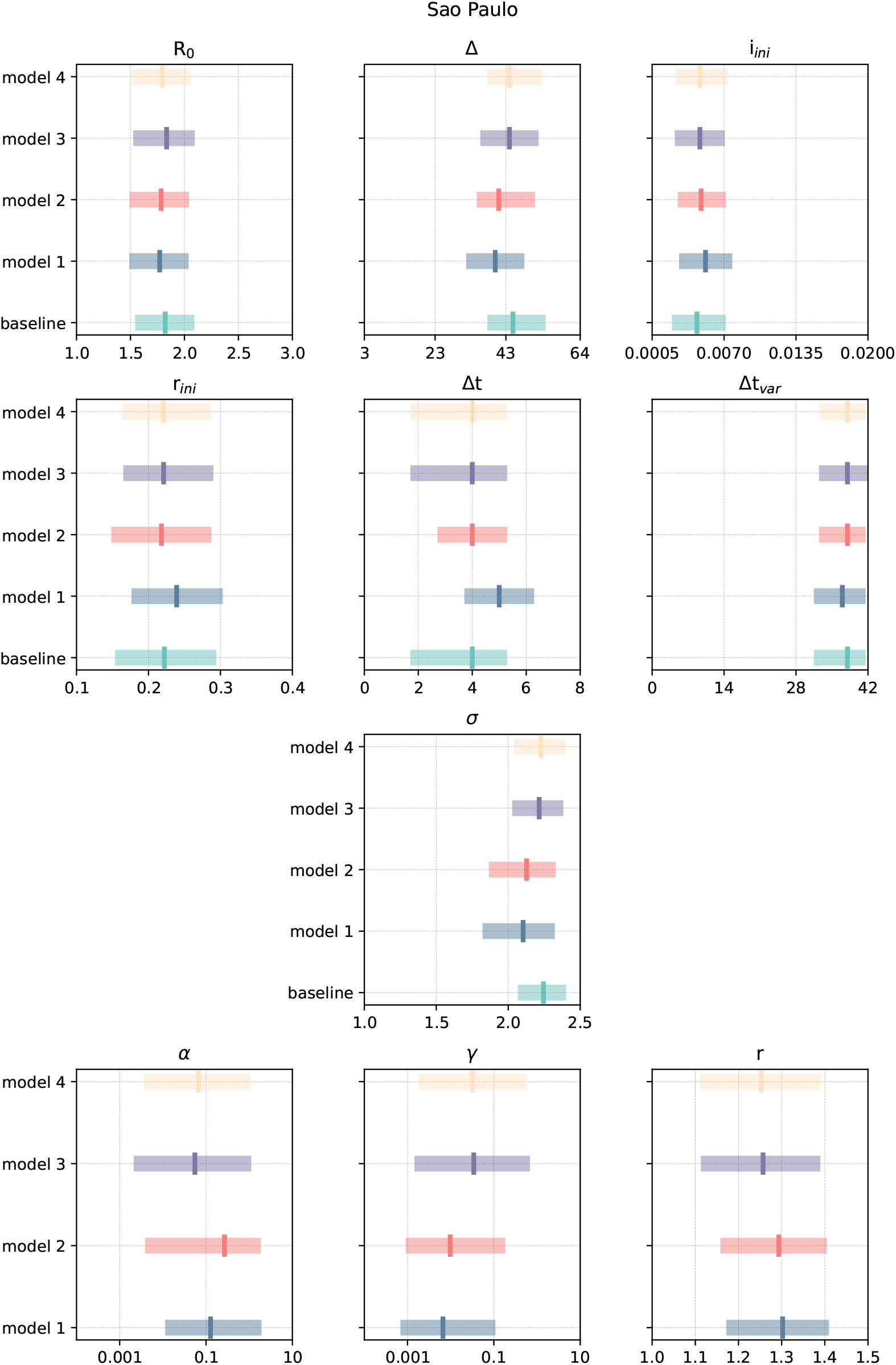
Posterior distributions of calibrated parameters for São Paulo. We plot the median with the range between the first quartile and third quartile. The first sixth parameters, *R*_0_, Δ, *i*_*ini*_, *r*_*ini*_, Δ*t*, Δ*t*_*var*_, *σ* are calibrated in all the five models. *α, β*, and *r* are behavioural parameters only in behavioural models (models 1-4).

Regarding the parameters in the behavioural models, parameters that control behavioural rate *α* and *γ* are sampled in a logarithm scale. We transform the sampled values by *exp*(*x*). The maximum values of medians of both *α* and *γ* are around 0.1 across the four regions. According to Fig., given *α*/*γ* equal to 0.1, the behavioural trasition rates from NC to C or from C to NC are under 0.2, exibiting a small behavioural values. This leads to the RDD/RDI results that behavioural mechanisms do not have a large impact on deaths and infections. Besides, Sao Paulo reports the largest *α* and the lowest *γ* compared to the other three regions, which is align with the largest RDD in Sao Paulo. The relative infection probability *r* of non-compliant individuals are similar across the regions within the range between 1.1 and 1.4, suggesting that non-compliant individuals have 1.1 to 1.4 times higher infection probablity than compliant individuals estimated by our models.

